# Comparison of NAFLD, MAFLD, MASLD and pure-MASLD characteristics and mortality outcomes in United States adults

**DOI:** 10.1101/2023.09.26.23296130

**Authors:** Rui Song, Zhao Li, Yingzhi Zhang, Jiahe Tan, Zhiwei Chen

## Abstract

**Background:** After metabolic dysfunction-associated fatty liver disease (MAFLD), non-alcoholic fatty liver disease (NAFLD) has recently been redefined again as metabolic dysfunction-associated steatotic liver disease (MASLD). However, the distinctions in characteristics and mortality outcomes between NAFLD, MAFLD, MASLD, and pure-MASLD are still unclear.

**Methods:** We analyzed data from 6,708 participants in the third National Health and Nutrition Examination Surveys 1988-1994 of United States (US) and their linked mortality through 2019. NAFLD, MAFLD, MASLD and pure MASLD were defined based on respective criteria. Survey-weight adjusted multivariable Cox proportional model was used to study the mortality of the four terms.

**Results:** The overall prevalence of NAFLD, MAFLD, MASLD and pure-MASLD was 28.9% (n=1,937), 26.1% (n=1,753), 29.0% (n=1,948), and 26.0% (n=1,741), respectively. For NAFLD, 81.3%, 89.9% and 89.9% fulfilled the criteria for MAFLD, MASLD and pure MASLD. For MAFLD, all were classified into the MASLD, and 89.8% were met the criteria of pure-MASLD. During a median follow-up of 27 years, both individuals with MAFLD and MASLD had higher risk of all-cause mortality (adjusted hazard ratio [aHR]: 1.21, 95% CI 1.09-1.352; and 1.11, 1.00-1.23, respectively). NAFLD and pure-MASLD were not associated with all-cause mortality. All the four terms were associated with increased all-cause mortality in individuals with advanced fibrosis (aHR: 1.66-1.83). Subgroup analyses showed that higher risk of all-cause mortality of NAFLD, MAFLD, MASLD and pure-MASLD were observed in female, age 41-55 years, non-Hispanic white, and never smoking subgroups when focused on moderate-severe hepatic steatosis.

**Conclusions:** In this US population-based study, MASLD could identify more individuals with all-cause mortality risk than MAFLD, meanwhile, NAFLD and pure-MASLD had similar characteristics and mortality outcomes.

## Introduction

Non-alcoholic fatty liver disease (NAFLD) has become one of the leading causes of chronic liver disease in the world, which can progress into hepatic fibrosis and end-stage liver diseases eventually ^1^. Most recently, a new term metabolic dysfunction-associated steatotic liver disease (MASLD) was recommended to replace NAFLD and metabolic dysfunction-associated fatty liver disease (MAFLD) by the Delphi consensus ^2^, which involved a change in not only the nomenclature but also the definition. MASLD, as the umbrella term, comprised broader metabolic risk factors, and can co-exist with other chronic liver diseases. To better distinguish the etiologies, MASLD was further classed into pure-MASLD, MetALD or MASLD with other combination etiology ^2^. Since the new term of MASLD was proposed, it is concern whether the new definition of MASLD will impact the old NAFLD data using, biomarker discovery or the nature history. Moreover, some are worried that this new term will add the difficulty and confusion to people due to frequent changes in nomenclature of NAFLD ^3 4^. Hence, it is urgent for us to clarify the relation characteristic and mortality outcomes of these terms in clinical practice.

Several studies found that NAFLD was highly in accordance with MASLD in clinical features ^5–8^, however, these studies were limited by the use of small or specific patient populations. A recent study reported similar prevalence and risk factors of NAFLD, MAFLD and MASLD in a large Brazilian cohort, but the long-term mortalities among these terms are not assessed ^9^. Some studies evaluated the mortality outcomes of MASLD and/or MAFLD, but showed inconsistent results ^10–12^. Meanwhile, the comparison of NAFLD, MAFLD, MASLD and pure-MASLD clinical outcomes was not performed in these previous studies. Therefore, the similarities and differences of characteristic and long-term mortality among individuals with NAFLD, MAFLD, MASLD and pure-MASLD in a general population are still unknown.

In this study, we conducted a comprehensive analysis in 6708 nonpregnant adult participants from the Third National Health and Nutrition Examination Survey 1988-1994 (NHANES III). By using the nationally representative sample of United States (US) adults, we compared the relationship and clinical features among NAFLD, MAFLD, MASLD and pure-MASLD, and assessed the association of the four terms on all-cause and cause-specific mortality.

## Materials and methods

### Study design and participants

The NHANES III is a national survey program to assess health and nutritional status obtained using a complex, stratified, clustered multi-staged probability sampling design to acquire a nationally representative sample of US populations ^13^. This survey included demographic data, health-related questionnaires, physical examinations, laboratory tests, and hepatic/gallbladder ultrasound in 13,588 nonpregnant adults (20-74 years).

First, we excluded 1,261 individuals with missing data on mortality status, body mass index, waist circumference, and laboratory tests, including high-density lipoprotein (HDL)-cholesterol, triglycerides, hemoglobin A1c and fasting glucose. Because of focusing on hepatic steatosis in this study, not steatohepatitis, we then excluded 1,223 individuals with elevated liver enzymes levels. To accurately assess the effect of alcohol intake on SLD, we further excluded 4,228 individuals with missing data on alcohol assumption. In addition, we also excluded 258 individuals with HCV and/or HBV infection, and those with a BMI <18.5 kg/m^2^, given the potential confounding effect of viral hepatitis and undernutrition. Lastly, 6,708 individuals were included for the main analysis in this study (Figure. S1). There is no significant difference of demographic and clinical characteristics among populations during selection process (Table S1). All individuals signed informed consent to participate in NHANES III.

### Data collection

For each participant of NHANES III, the information about demographic indexes (such as sex, age, ethnicity, education, marital status, smoking, alcohol consumption, physical activity, underlying diseases, and drug use), anthropometric indexes (such as body mass index (BMI), waist circumference, and blood pressure), and laboratory tests (such as alanine aminotransferase, aspartate aminotransferase, HDL-cholesterol, fasting glucose) were collected. The definitions, such as hypertension, diabetes, smoking status, alcohol consumption, sedentary behavior, were provided in the supplementary materials.

### Definition of NAFLD, MAFLD, MASLD, pure-MASLD and advanced fibrosis

In NHANES III, the ultrasonographic assessments were reported as normal vs. mild, moderate, or severe hepatic steatosis ^14^. We defined steatosis liver diseases (SLD) as mild to severe hepatic steatosis without increased liver enzymes levels in this study. NAFLD was defined as the presence of SLD without excessive alcohol consumption (≥20 g/d for females and ≥30 g/d for males) and/or other liver diseases (such as viral hepatitis) ^15^. MAFLD was defined as the presence of SLD with metabolic dysfunction, which comprises either overweight or obese (BMI ≥25 kg/m^2^), diabetes mellitus, or a combination of at least 2 of the 7 metabolic risk abnormalities ^16^. MASLD was defined as the presence of SLD combined with at least 1 of the 5 cardiometabolic adult criteria ^2^. Pure-MASLD was defined as the presence of MASLD without excessive alcohol consumption and other combination etiologies ^2^. MetALD was defined as the presence of MASLD with higher alcohol consumption levels (20 to 50 g/d for females and 30 to 60 g/d for males) ^2^. This detail definitions of MAFLD and MASLD were showed in the supplementary materials.

To give an insight into the similarities and differences between the four terms, individuals were further classified into different groups. For example, we defined NAFLD+ as individuals who met the definition of NAFLD, and NAFLD- was defined as individuals who didn’t meet NAFLD definition. Likewise, NAFLD+/MAFLD-/MASLD-/pure-MASLD+ means an individual who met both NAFLD and pure-MASLD definitions but not the definitions of MAFLD and MASLD. Individuals without hepatic steatosis were referred to as a group with no hepatic steatosis.

We using the NAFLD fibrosis score (NFS) to evaluate advanced fibrosis of individual with SLD^17^. In detail, high (NFS >0.676), intermediate (NFS 0.676 to -1.455) and low probability for advanced fibrosis (NFS <-1.455) were defined.

### All-cause and cause-specific mortality

The mortality data of adults in the NHANES III was passively follow-up and obtained from the National Death Index, which contained complete data until December 31, 2019 ^18^. The definition of all-cause mortality and cause-specific (cardiovascular disease and cancer) mortalities were showed in a previous study ^19^, and detailed information was presented in the supplementary materials. Moreover, we calculated the follow-up time (month) from the date of interview to the date of death or the end of the follow-up period.

### Statistical analysis

The results of the baseline characteristics are expressed as median and interquartile range (IQR) for continuous variables and unweighted frequency counts and weighted percentage for categorical variables. We compared baseline characteristics using the Kruskal-Wallis test for continuous variables and the chi-square test for categorical variables. Cox proportional hazards regression was used to estimate hazard ratios (HR) and 95% confidence intervals (CI) for deaths from all-causes, cardiovascular disease and cancer. To avoid an overadjustment bias in the multivariable analysis ^20^, we discarded the related variables which have presented in the definition of NAFLD, MAFLD and MASLD. We used two models with progressive degrees of adjustment: model 1 adjusted for age, sex, and ethnicity; model 2 further adjusted for education, marital status, smoking status, and sedentary lifestyle.

For sensitivity analysis, first, we added participants with hepatitis B, hepatitis C, or BMI < 18.5 kg/m^2^ into the entire population. Second, to assess the effect of varying degrees of SLD, we carried out another sensitivity analysis comparing the group with no hepatic steatosis against those with moderate-severe hepatic steatosis (that is, excluding those with mild steatosis from individuals with SLD).

Lastly, we evaluated the association of NAFLD, MAFLD, MASLD and pure-MASLD with all-cause mortality for subgroups defined by age, sex, ethnicity, BMI, diabetes, hypertension, and smoking status. All analyses were performed with R (version 4.3.0; R Foundation for Statistical Computing, Vienna, Austria), using the survey package to account for the sampling weights and the complex survey design in the NHANES III.

## Results

### Characteristics of NAFLD, MAFLD, MASLD and pure-MASLD

After included individuals with available alcohol consumption and liver ultrasound data and excluded individuals with increased liver enzymes levels or viral hepatitis etc., a total of 6708 participants from NHANES III were eligible for the final analysis (Figure. S1). The overall prevalence of SLD, NAFLD, MAFLD, MASLD and pure-MASLD was 32.3% (n=2,164), 28.9% (n=1,937), 26.1% (n=1,753), 29.0% (n=1948), and 26.0% (n=1,741), respectively (Figure 1). Compared no hepatic steatosis group, older, less educated, obese, sedentary, and more coexisting metabolic risk factors were observed in SLD group. Among NAFLD, MAFLD, MASLD and pure-MASLD groups, individuals with MAFLD were more likely to be older, male, higher BMI and waist circumference, higher alcohol intake, hypertension, and diabetes. (Table S2)

**Figure 1.**
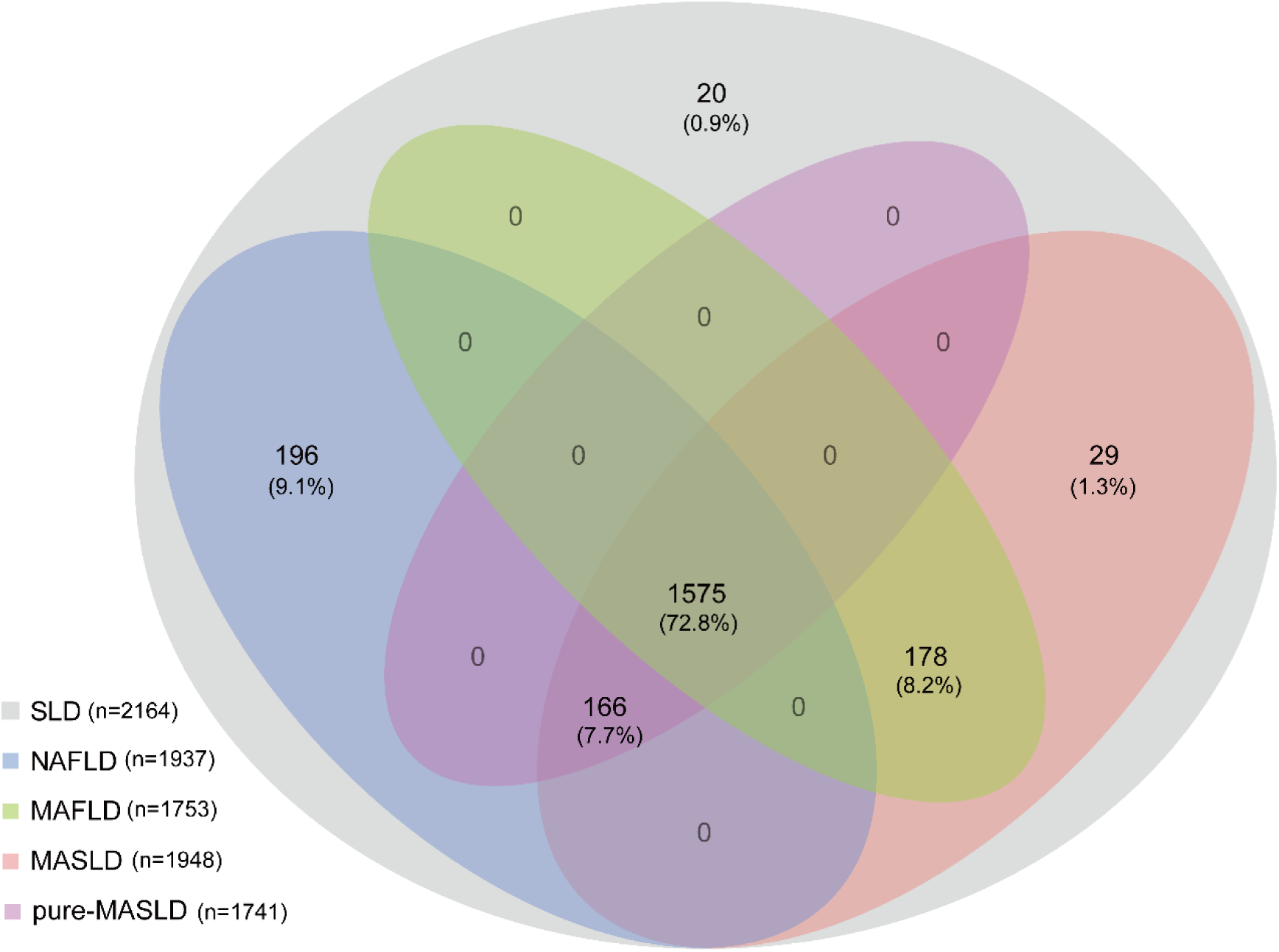
Venn diagram of the relationship among NAFLD, MAFLD, MASLD and pure-MASLD. SLD, steatotic liver disease; MAFLD, dysfunction-associated fatty liver disease; MASLD, metabolic dysfunction-associated steatotic liver disease; NAFLD, non-alcoholic fatty liver disease metabolic.

Further classified into different combinations of the 2,164 SLD individuals, six groups were generated (NAFLD+/MAFLD+/MASLD+/pure-MASLD+, NAFLD+/MAFLD-/MASLD+/pure- MASLD+, NAFLD-/MAFLD+/MASLD+/pure-MASLD-, NAFLD+/MAFLD-/MASLD-/pure- MASLD-, NAFLD-/MAFLD-/MASLD+/pure-MASLD-, and NAFLD-/MAFLD-/MASLD-/pure-MASLD-; Figure 1). As shown in Figure 1, most individuals with SLD were classified into NAFLD+/MAFLD+/MASLD+/pure-MASLD+ (72.8%), followed by NAFLD+/MAFLD-/MASLD-/pure-MASLD- (9.1%), NAFLD-/MAFLD+/MASLD+/pure-MASLD- (8.2%), NAFLD+/MAFLD-/MASLD+/pure-MASLD+ (7.7%), NAFLD-/MAFLD-/MASLD+/pure- MASLD- (1.3%), and NAFLD-/MAFLD-/MASLD-/pure-MASLD- (0.9%). For individuals with NAFLD, 89.9% individuals were in accordance with MASLD and pure-MASLD, and 81.3% individuals were in accordance with MAFLD. For individuals with MAFLD, 89.8% individuals were in accordance with NAFLD and pure-MASLD, and 100% individuals in accordance with MASLD. For individuals with MASLD, 89.4% individuals were in accordance with NAFLD and pure-MASLD, and 90.0% individuals were in accordance with MAFLD. For individuals with pure-MASLD, 100% individuals were in accordance with NAFLD and MASLD, and 90.5% individuals were in accordance with MAFLD. The baseline characteristics of different combinations of SLD individuals were presented in Table 1. Similarly, older, obese, hypertension, diabetes, and more metabolic risk factors were more presented in individuals with MAFLD+. In addition, men, and higher alcohol intake were more observed in individuals with NAFLD-. Individuals with NAFLD+MAFLD- were more likely to be younger, female, lower BMI and waist circumference, and lower metabolic risk factors.

**Table 1.** Baseline characteristics of the study population (N = 6,708).

| **Characteristics** | **No hepatic**<br>**steatosis**<br>**(n=4,544)** | **Group 1<sup>#</sup>**<br>**(n=1,575)** | **Group 2<sup>#</sup>**<br>**(n=178)** | **Group 3<sup>#</sup>**<br>**(n=166)** | **Group 4<sup>#</sup>**<br>**(n=196)** | **Group 5<sup>#</sup>**<br>**(n=29)** | **Group 6<sup>#</sup>**<br>**(n=20)** | **P** |
| --- | --- | --- | --- | --- | --- | --- | --- | --- |
| Age (years) | 36.0 (28.0, 48.0) | 47.0 (36.0, 60.0) | 47.0 (35.0, 58.9) | 32.0 (26.0, 44.0) | 30.0 (25.0, 39.0) | 37.0 (29.3, 62.0) | 30.0 (27.2, 31.5) | <0.001 |
| Sex (male) | 2,301 (52%) | 773 (56%) | 135 (76%) | 57 (30%) | 86 (41%) | 23 (76%) | 15 (69%) | <0.001 |
| Race/ethnicity |  |  |  |  |  |  |  | 0.10 |
| Non-Hispanic white | 1,814 (78%) | 597 (75%) | 76 (86%) | 75 (83%) | 80 (82%) | 12 (81%) | 9 (83%) |  |
| Non-Hispanic black | 1,198 (4.5%) | 560 (6.3%) | 62 (5.1%) | 43 (3.5%) | 50 (4.7%) | 8 (5.3%) | 2 (1.2%) |  |
| Mexican-American | 1,324 (9.8%) | 348 (8.2%) | 36 (6.3%) | 40 (7.8%) | 61 (8.4%) | 9 (13%) | 8 (8.7%) |  |
| Others | 208 (8.1%) | 70 (10%) | 4 (2.9%) | 8 (5.9%) | 5 (4.4%) | 0 (0%) | 1 (7.0%) |  |
| Body mass index (kg/m<sup>2</sup>) | 24.6 (22.3, 27.3) | 29.3 (26.5, 32.4) | 28.3 (25.9, 32.2) | 22.0 (20.9, 23.7) | 21.7 (20.4, 22.9) | 22.7 (20.6, 24.1) | 21.2 (19.7, 21.9) | <0.001 |
| Waist circumference (cm) | 88 (79, 96) | 102 (94, 110) | 100 (91, 110) | 81 (73, 85) | 74 (70, 79) | 84 (81, 91) | 75 (70, 79) | <0.001 |
| ≥12 years education | 3,094 (82%) | 874 (73%) | 106 (78%) | 111 (82%) | 138 (81%) | 18 (86%) | 12 (61%) | <0.001 |
| Marital status | 2,798 (65%) | 1,097 (75%) | 117 (64%) | 101 (62%) | 117 (73%) | 17 (81%) | 10 (63%) | <0.001 |
| Hypertension | 1,140 (20%) | 691 (43%) | 105 (67%) | 22 (8.9%) | 5 (2.7%) | 9 (28%) | 2 (10%) | <0.001 |
| Diabetes | 219 (2.8%) | 298 (13%) | 25 (9.1%) | 1 (0.8%) | 0 (0%) | 0 (0%) | 0 (0%) | <0.001 |
| Smoking status |  |  |  |  |  |  |  | <0.001 |
| Never | 2,297 (46%) | 826 (48%) | 41 (30%) | 91 (49%) | 111 (57%) | 6 (11%) | 5 (15%) |  |
| Past smoker | 866 (22%) | 427 (30%) | 64 (37%) | 24 (15%) | 22 (14%) | 6 (23%) | 2 (16%) |  |
| Current smoker | 1,381 (32%) | 322 (22%) | 73 (33%) | 51 (36%) | 63 (29%) | 17 (66%) | 13 (68%) |  |
| Alcohol consumption (g/d) | 4 (1, 12) | 2 (0, 8) | 42 (36, 60) | 3 (1, 6) | 4 (1, 12) | 48 (32, 56) | 61 (36, 82) | <0.001 |
| Sedentary lifestyle | 1,061 (16%) | 520 (24%) | 42 (16%) | 41 (17%) | 37 (9.5%) | 9 (28%) | 6 (42%) | <0.001 |
| Triglycerides (mg/dL) | 1.1 (0.8, 1.6) | 1.8 (1.2, 2.6) | 1.6 (1.2, 2.3) | 1.0 (0.8, 1.3) | 0.9 (0.7, 1.2) | 0.8 (0.7, 1.1) | 0.7 (0.5, 1.1) | <0.001 |
| HDL-cholesterol (mg/dL) | 1.3 (1.1, 1.6) | 1.1 (0.9, 1.3) | 1.3 (1.0, 1.5) | 1.3 (1.1, 1.5) | 1.6 (1.3, 1.8) | 1.5 (1.3, 1.8) | 1.9 (1.3, 2.3) | <0.001 |
| C-reactive protein (mg/dL) | 0.2 (0.2, 0.2) | 0.2 (0.2, 0.5) | 0.2 (0.2, 0.4) | 0.2 (0.2, 0.2) | 0.2 (0.2, 0.2) | 0.2 (0.2, 0.2) | 0.2 (0.2, 0.2) | <0.001 |
| Glucose (mol/L) | 5.1 (4.8, 5.4) | 5.4 (5.0, 5.8) | 5.3 (4.9, 5.6) | 5.0 (4.7, 5.3) | 4.9 (4.7, 5.2) | 5.2 (4.9, 5.5) | 5.0 (4.5, 5.6) | <0.001 |
| Insulin (uU/mL) | 7.0 (5.4, 9.7) | 11.4 (8.3, 15.9) | 9.6 (7.1, 13.6) | 7.1 (5.1, 8.5) | 5.6 (4.7, 7.4) | 5.3 (4.1, 6.9) | 4.2 (4.2, 5.4) | <0.001 |
| Hemoglobin A1c (%) | 5.1 (4.8, 5.4) | 5.4 (5.0, 5.7) | 5.2 (5.0, 5.5) | 5.1 (4.8, 5.3) | 5.0 (4.6, 5.3) | 5.0 (4.5, 5.3) | 5.1 (4.7, 5.1) | <0.001 |
| HOMA-IR | 1.6 (1.2, 2.3) | 2.8 (2.0, 4.2) | 2.2 (1.6, 3.3) | 1.5 (1.1, 1.9) | 1.2 (1.0, 1.7) | 1.2 (1.0, 1.5) | 1.1 (0.9, 1.3) | <0.001 |

Data are shown as the median (interquartile range) or unweighted frequency counts (weighted percentage) as appropriate. The Kruskal-Wallis test for continuous variables and the Chi-square test for categorical variables were used in this analysis.

### All-cause and cause-specific mortality of NAFLD, MAFLD, MASLD and pure-MASLD

During the median follow-up of 27.0 (interquartile range: 24.6-28.9) years, the cumulative mortality from all causes was 30.7% (2,061 deaths). For cause-specific mortality, cardiovascular disease was 537 deaths (8.0%), and cancer was 523 deaths (7.8%). In total, individuals with SLD had a 1.12-fold higher all-cause mortality than those without SLD after adjustment for sociodemographic characteristics and lifestyle risk factors (model 2: adjusted HR (aHR): 1.12, 95% CI 1.00-1.25, Table 2). In detail, individuals with MAFLD showed a higher all-cause mortality than those without MAFLD in multivariable model 2 (aHR: 1.21, 95% CI 1.09-1.35). Similar result was also observed in MASLD individuals (aHR: 1.11, 95% CI 1.00-1.23). But the increased risks of all-cause mortality were not presented in individuals with NAFLD or pure-MASLD (aHR: 1.07, 95% CI 0.95-1.20; aHR: 1.07, 95% CI 0.96-1.19, respectively). (Table 2) After stratified by etiology of MASLD, we found that only MASLD with ALD (>50 g/d for females and >60 g/d for males) was associated with increased all-cause mortality compared with individuals with no hepatic steatosis (aHR: 2.10, 95% CI 1.22-3.60). Individuals with MetALD were not associated with all-cause mortality (aHR:1.17, 95% CI 0.84-1.64). (Table S3)

**Table 2.**
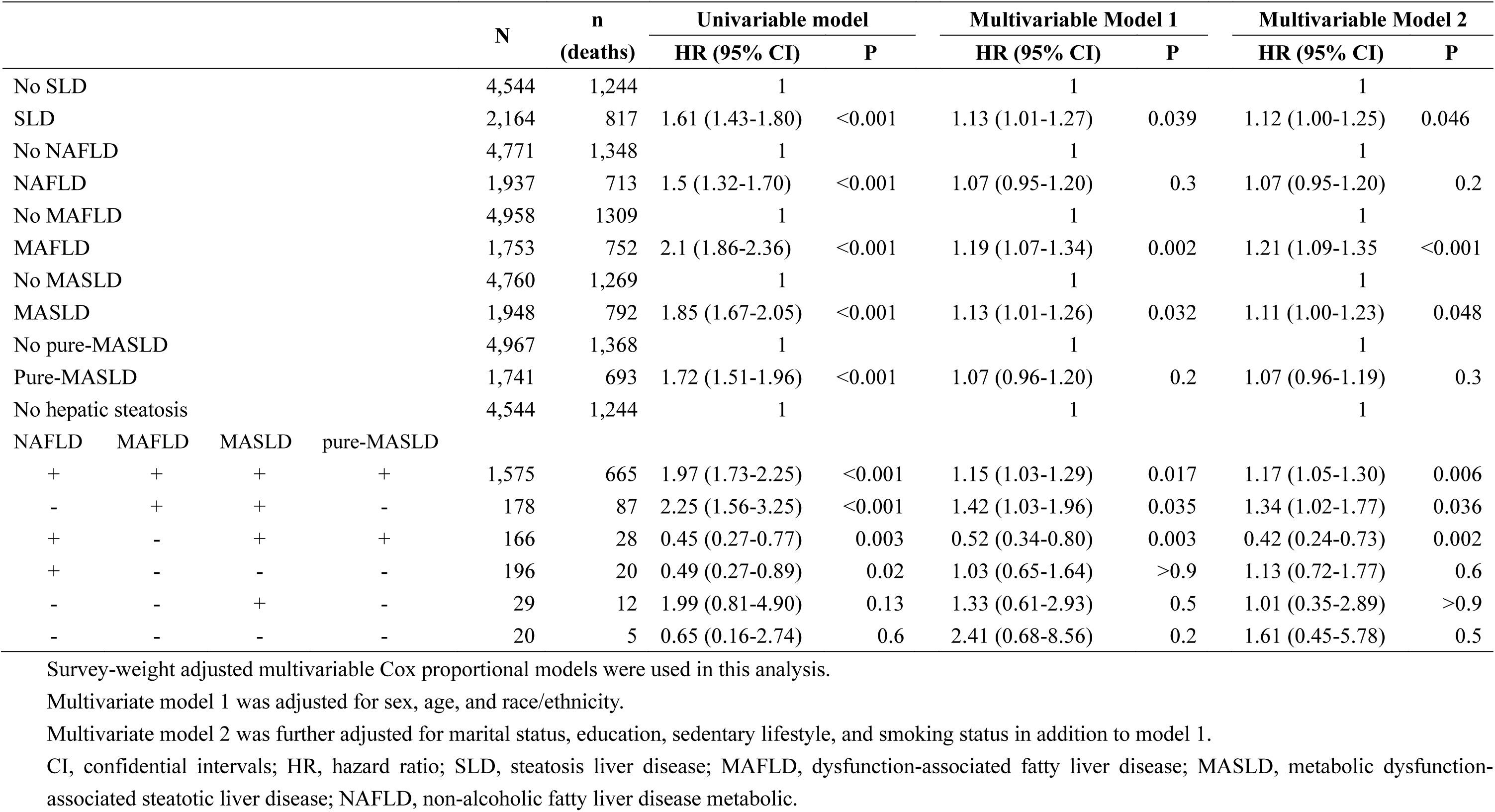
Association among NAFLD, MAFLD, MASLD and pure-MASLD status, and all-cause mortality.

To further clarify the all-cause mortality among NAFLD, MAFLD, MASLD and pure-MASLD, we classified SLD individuals into six groups as mentioned above. As shown in Table 2, individuals with NAFLD-/MAFLD+/MASLD+/pure-MASLD- had the highest risk of all-cause mortality in multivariable model 2 (aHR: 1.34, 95% CI 1.02-1.77), followed by those with NAFLD+/MAFLD+/MASLD+/pure-MASLD+ (aHR: 1.17, 95% CI 1.05-1.30). Unexpectedly, individuals with NAFLD+/MAFLD-/MASLD+/pure-MASLD+ showed a lower risk of all-cause mortality compared with individuals with no hepatic steatosis (aHR: 0.42, 95% CI 0.24-0.73). The rest of groups were not associated with all-cause mortality.

In subgroup analyses, MAFLD was associated with increased all-cause mortality in female, age >40 years, non-Hispanic white, BMI 18.5-24.9 kg/m^2^, without diabetes and current smoking subgroups (aHR: 1.16-1.57, Figure 2A). For MASLD, similar results were only observed in age 41-55 years and non-Hispanic white subgroups. Pure-MASLD was only associated with a higher risk of all-cause mortality in age 41-55 years subgroup. And NAFLD was not associated with all-cause mortality in any subgroups. (Figure 2A)

**Figure 2.**
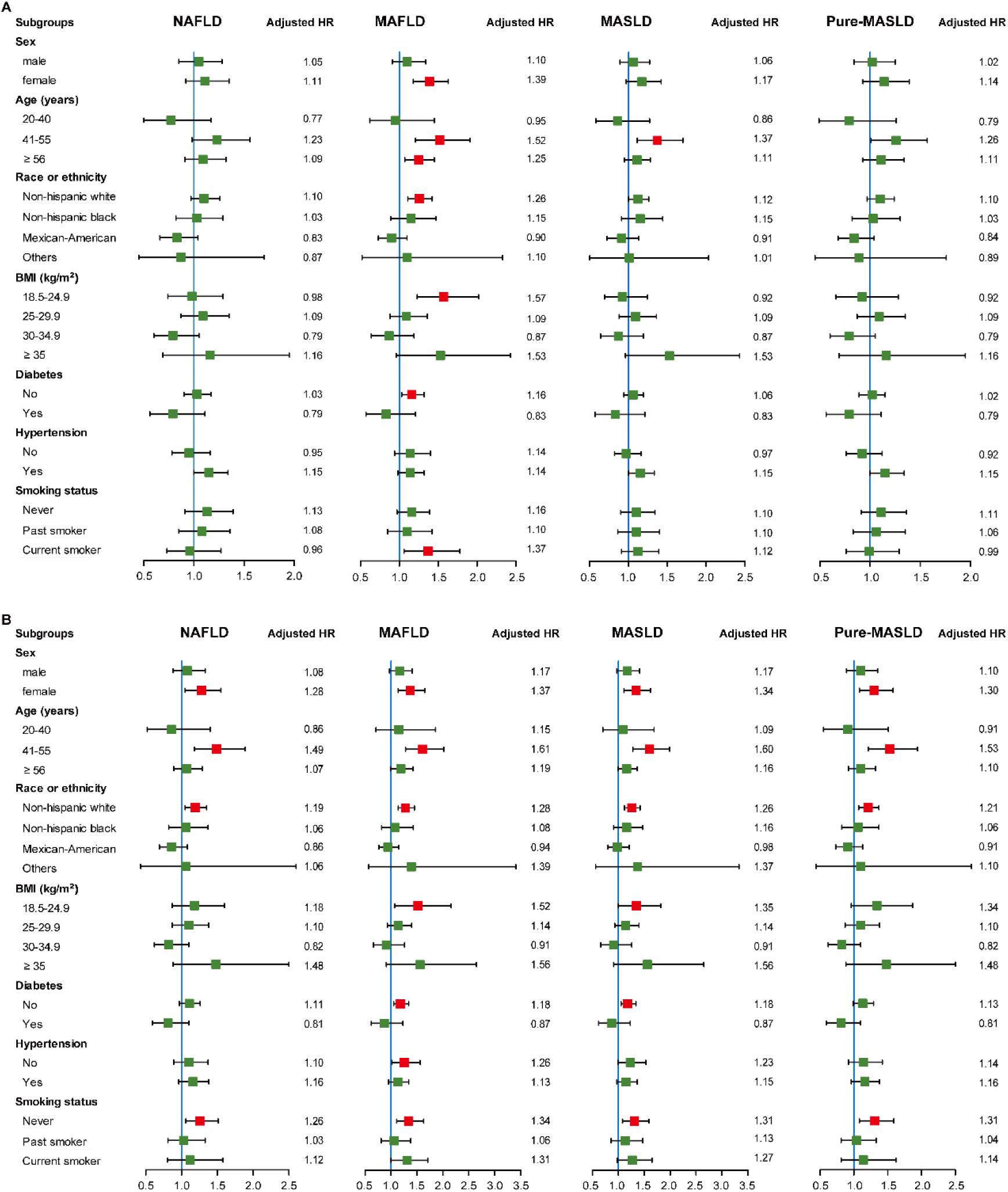
Subgroup analyses of association with all-cause mortality among NAFLD, MAFLD, MASLD and pure-MASLD in total SLD individuals (A) and in moderate-severe SLD individuals (B). Green box means HR value without statistical difference. Red box means HR value with statistical difference. The bars on both sides of box mean 95% CI of HR. The adjusted model 2 (sex, age, race/ethnicity, marital status, education, sedentary lifestyle, and smoking status) was used in this analysis. BMI, body mass index; CI, confidence intervals. HR, hazard ratio; SLD, steatotic liver disease; MAFLD, dysfunction-associated fatty liver disease; MASLD, metabolic dysfunction-associated steatotic liver disease; NAFLD, non-alcoholic fatty liver disease metabolic.

When the analysis was restricted to cause-specific mortality, all the four terms and their combinations were not showed a significant association with cardiovascular mortality or cancer mortality (Table 3). But one combination was excepted, that is, individuals with NAFLD+/MAFLD-/MASLD+/pure-MASLD+ showed both lower risk of cardiovascular mortality and cancer mortality (aHR: 0.17, 95% CI 0.03-0.83; aHR: 0.22, 95% CI 0.07-0.64, respectively). Lastly, we evaluated the effect of fibrosis on the all-cause mortality among NAFLD, MAFLD, MASLD and pure-MASLD. As shown in Table 4, compared to low NFS score group, all the four terms showed increased risk of all-cause mortality irrespective in intermediate NFS score group or high NFS score group, which was larger in high NFS score group (intermediate NFS: aHR: 1.27-1.53; high NFS: aHR: 1.66-1.83, respectively).

**Table 3.** Association among NAFLD, MAFLD, MASLD and pure-MASLD status, and cardiovascular disease and cancer-related mortality.

|  |  |  |  | N | n<br>(deaths) | Univariable model |  | Multivariable Model 1 |  | Multivariable Model 2 |  |
| --- | --- | --- | --- | --- | --- | --- | --- | --- | --- | --- | --- |
|  |  |  |  |  |  | HR (95% CI) | P | HR (95% CI) | P | HR (95% CI) | P |
| Cardiovascular mortality |  |  |  |  |  |  |  |  |  |  |  |
| No NAFLD |  |  |  | 4,771 | 347 | 1 |  | 1 |  | 1 |  |
| NAFLD |  |  |  | 1,937 | 190 | 1.43 (1.07-1.93) | 0.017 | 0.97 (0.73-1.29) | 0.8 | 0.96 (0.71-1.30) | 0.8 |
| No MAFLD |  |  |  | 4,958 | 332 | 1 |  | 1 |  | 1 |  |
| MAFLD |  |  |  | 1,753 | 205 | 2.23 (1.71-2.91) | <0.001 | 1.19 (0.92-1.53) | 0.2 | 1.21 (0.94-1.57) | 0.14 |
| No MASLD |  |  |  | 4,760 | 324 | 1 |  | 1 |  | 1 |  |
| MASLD |  |  |  | 1,948 | 213 | 1.90 (1.44-2.49) | <0.001 | 1.08 (0.83-1.42) | 0.5 | 1.06 (0.81-1.40) | 0.7 |
| No pure-MASLD |  |  |  | 4,967 | 350 | 1 |  | 1 |  | 1 |  |
| Pure-MASLD |  |  |  | 1,741 | 187 | 1.74 (1.31-2.30) | <0.001 | 1.01 (0.76-1.35) | >0.9 | 1.00 (0.74-1.36) | >0.9 |
| No hepatic steatosis |  |  |  | 4,544 | 319 | 1 |  | 1 |  | 1 |  |
| NAFLD | MAFLD | MASLD | pure-MASLD |  |  |  |  |  |  |  |  |
| + | + | + | + | 1,575 | 182 | 2.02 (1.52-2.67) | <0.001 | 1.1 (0.83- 1.46) | 0.5 | 1.12 (0.84, 1.48) | 0.4 |
| - | + | + | - | 178 | 23 | 2.53 (1.37-4.67) | 0.003 | 1.55 (0.79- 3.04) | 0.2 | 1.47 (0.79, 2.74) | 0.2 |
| + | - | + | + | 166 | 5 | 0.20 (0.04-0.96) | 0.015 | 0.23 (0.05- 1.07) | 0.06 | 0.17 (0.03, 0.83) | 0.029 |
| + | - | - | - | 196 | 3 | 0.11 (0.02-0.67) | 0.016 | 0.27 (0.04- 1.70) | 0.2 | 0.31 (0.06, 1.69) | 0.2 |
| - | - | + | - | 29 | 3 | 1.20 (0.27-5.34) | 0.8 | 0.71 (0.14- 3.71) | 0.7 | 0.63 (0.13, 3.11) | 0.6 |
| - | - | - | - | 20 | 2 | 1.53 (0.22-10.80) | 0.7 | 8 (1.44- 44.4) | 0.017 | 4.99 (0.88, 28.2) | 0.069 |
| Cancer mortality |  |  |  |  |  |  |  |  |  |  |  |
| No NAFLD |  |  |  | 4,771 | 358 | 1 |  | 1 |  | 1 |  |
| NAFLD |  |  |  | 1,937 | 165 | 1.27 (0.93, 1.74) | 0.14 | 0.94 (0.69, 1.29) | 0.7 | 0.99 (0.72, 1.36) | >0.9 |
| No MAFLD |  |  |  | 4,958 | 348 | 1 |  | 1 |  | 1 |  |
| MAFLD |  |  |  | 1,753 | 175 | 1.89 (1.46, 2.45) | <0.001 | 1.13 (0.86, 1.48) | 0.4 | 1.21 (0.92, 1.60) | 0.2 |
| No MASLD |  |  |  | 4,760 | 337 | 1 |  | 1 |  | 1 |  |
| MASLD |  |  |  | 1,948 | 186 | 1.63 (1.29, 2.07) | <0.001 | 1.04 (0.80, 1.35) | 0.8 | 1.07 (0.83, 1.39) | 0.6 |
| No pure-MASLD |  |  |  | 4,967 | 363 | 1 |  | 1 |  | 1 |  |
| Pure-MASLD |  |  |  | 1,741 | 160 | 1.44 (1.07, 1.94) | 0.016 | 0.94 (0.70, 1.27) | 0.7 | 0.98 (0.73, 1.33) | >0.9 |
| No hepatic steatosis |  |  |  | 4,544 | 331 | 1 |  | 1 |  | 1 |  |
| NAFLD | MAFLD | MASLD | pure-MASLD |  |  |  |  |  |  |  |  |
| + | + | + | + | 1,575 | 152 | 1.69 (1.24- 2.32) | 0.001 | 1.04 (0.76- 1.41) | 0.8 | 1.11 (0.81- 1.52) | 0.5 |
| - | + | + | - | 178 | 23 | 2.59 (1.44- 4.67) | 0.001 | 1.63 (0.92- 2.87) | 0.092 | 1.61 (0.93- 2.78) | 0.09 |
| + | - | + | + | 166 | 8 | 0.26 (0.10- 0.64) | 0.004 | 0.3 (0.12- 0.77) | 0.012 | 0.22 (0.07- 0.64) | 0.006 |
| + | - | - | - | 196 | 5 | 0.52 (0.15- 1.75) | 0.3 | 0.99 (0.30- 3.26) | >0.9 | 1.07 (0.31- 3.66) | >0.9 |
| - | - | + | - | 29 | 3 | 1.69 (0.39- 7.27) | 0.5 | 1.19 (0.23- 6.09) | 0.8 | 0.8 (0.14- 4.47) | 0.8 |
| - | - | - | - | 20 | 1 | 0.74 (0.09- 6.32) | 0.8 | 1.87 (0.23- 15.1) | 0.6 | 1.21 (0.14- 10.2) | 0.9 |

Survey-weight adjusted multivariable Cox proportional models were used in this analysis.
Multivariate model 1 was adjusted for sex, age, and race/ethnicity.
Multivariate model 2 was further adjusted for marital status, education, sedentary lifestyle, and smoking status in addition to model 1.
CI, confidential intervals; HR, hazard ratio; MAFLD, dysfunction-associated fatty liver disease; MASLD, metabolic dysfunction-associated steatotic liver disease; NAFLD, non-alcoholic fatty liver disease metabolic.

**Table 4.** Association of advanced fibrosis status and all-cause mortality among individuals with NAFLD, MAFLD, MASLD and pure-MASLD.

|  | N | n<br>(deaths) | Univariable model |  | Multivariable Model 1 |  | Multivariable Model 2 |  |
| --- | --- | --- | --- | --- | --- | --- | --- | --- |
|  |  |  | HR (95% CI) | P | HR (95% CI) | P | HR (95% CI) | P |
| NAFLD |  |  |  |  |  |  |  |  |
| Low NFS | 1,277 | 294 | 1 |  | 1 |  | 1 |  |
| Intermediate NFS | 545 | 331 | 4.07 (3.18, 5.21) | <0.001 | 1.24 (1.00, 1.53) | 0.048 | 1.27 (1.04, 1.55) | 0.02 |
| High NFS | 97 | 79 | 6.98 (4.44, 11.0) | <0.001 | 1.87 (1.25, 2.80) | 0.002 | 1.79 (1.24, 2.60) | 0.002 |
| MAFLD |  |  |  |  |  |  |  |  |
| Low NFS | 1,064 | 299 | 1 |  | 1 |  | 1 |  |
| Intermediate NFS | 570 | 359 | 3.74 (2.90, 4.81) | <0.001 | 1.45 (1.16, 1.80) | 0.001 | 1.53 (1.24, 1.89) | <0.001 |
| High NFS | 103 | 85 | 6.35 (4.16, 9.68) | <0.001 | 1.93 (1.32, 2.81) | <0.001 | 1.83 (1.29, 2.59) | <0.001 |
| MASLD |  |  |  |  |  |  |  |  |
| Low NFS | 1,227 | 323 | 1 |  | 1 |  | 1 |  |
| Intermediate NFS | 595 | 373 | 3.9 (3.04, 5.00) | <0.001 | 1.38 (1.12, 1.70) | 0.003 | 1.41 (1.15, 1.72) | <0.001 |
| High NFS | 106 | 86 | 6.34 (4.15, 9.69) | <0.001 | 1.81 (1.25, 2.63) | 0.002 | 1.66 (1.14, 2.42) | 0.008 |
| Pure-MASLD |  |  |  |  |  |  |  |  |
| Low NFS | 1,097 | 280 | 1 |  | 1 |  | 1 |  |
| Intermediate NFS | 532 | 327 | 3.79 (3.00, 4.81) | <0.001 | 1.28 (1.03, 1.58) | 0.027 | 1.31 (1.06, 1.62) | 0.011 |
| High NFS | 95 | 78 | 6.73 (4.28, 10.6) | <0.001 | 1.88 (1.22, 2.91) | 0.005 | 1.8 (1.20, 2.69) | 0.004 |

Survey-weight adjusted multivariable Cox proportional models were used in this analysis.
Multivariate model 1 was adjusted for sex, age, and race/ethnicity.
Multivariate model 2 was further adjusted for marital status, education, sedentary lifestyle, and smoking status in addition to model 1.
CI, confidential intervals; HR, hazard ratio; MAFLD, dysfunction-associated fatty liver disease; MASLD, metabolic dysfunction-associated steatotic liver disease;
NAFLD, non-alcoholic fatty liver disease metabolic; NFS, NAFLD fibrosis score.

### Sensitivity analyses of NAFLD, MAFLD, MASLD and pure-MASLD

Firstly, we want to know if other etiologies of SLD will affect the clinical outcomes. After added individuals with other confounders (viral hepatitis and low BMI) to the entire population, similar findings of mortality outcomes were observed (Table S4-7). And then we wonder whether the extent of hepatic steatosis may have an influence on the mortality. After classified into three groups (mild, moderate, and severe) of SLD according to the ultrasound results in NHANES III, we found individuals with moderate and severe SLD were significantly associated with increased all-cause mortality, and had similar effect (aHR: 1.21, 95% CI 1.06-1.39; aHR: 1.22, 95% CI 1.03-1.45, respectively; Table S8). Hence, we conducted another sensitivity analysis excluding the mild hepatic steatosis from SLD (that is, SLD were redefined as those with moderate-severe hepatic steatosis). Notably, in this scenario, not only MAFLD and MASLD were associated with increased all-cause mortality (aHR: 1.25, 95% CI 1.13-1.39; aHR: 1.24, 95% CI 1.12-1.38, respectively), but also NAFLD and pure-MASLD did (aHR: 1.16, 95% CI 1.04-1.30; aHR: 1.18, 95% CI 1.05-1.33, respectively; Table 5). Regarding the different of combinations, the results of NAFLD+/MAFLD+/MASLD+/pure-MASLD+ group and NAFLD-/MAFLD+/MASLD+/pure- MASLD- group were in line with our previous findings (aHR: 1.23, 95% CI 1.10-1.38; aHR: 1.39, 95% CI 1.06-1.83, respectively). However, the lower risk of all-cause mortality of NAFLD+/MAFLD-/MASLD+/pure-MASLD+ group in previous was disappeared in this sensitivity analysis, instead of the NAFLD-/MAFLD-/MASLD+/pure-MASLD- group had a higher risk of all-cause mortality (aHR: 2.65, 95% CI 1.70-4.12). (Table 5) Compared to no hepatic steatosis, individuals with MetALD had a higher risk of all-cause mortality (aHR: 1.41, 95% CI 1.02-1.94), whereas MASLD with ALD was not associated with all-cause mortality (aHR: 1.95, 95% CI 0.97-3.91, P=0.06; Table S9).

**Table 5.**
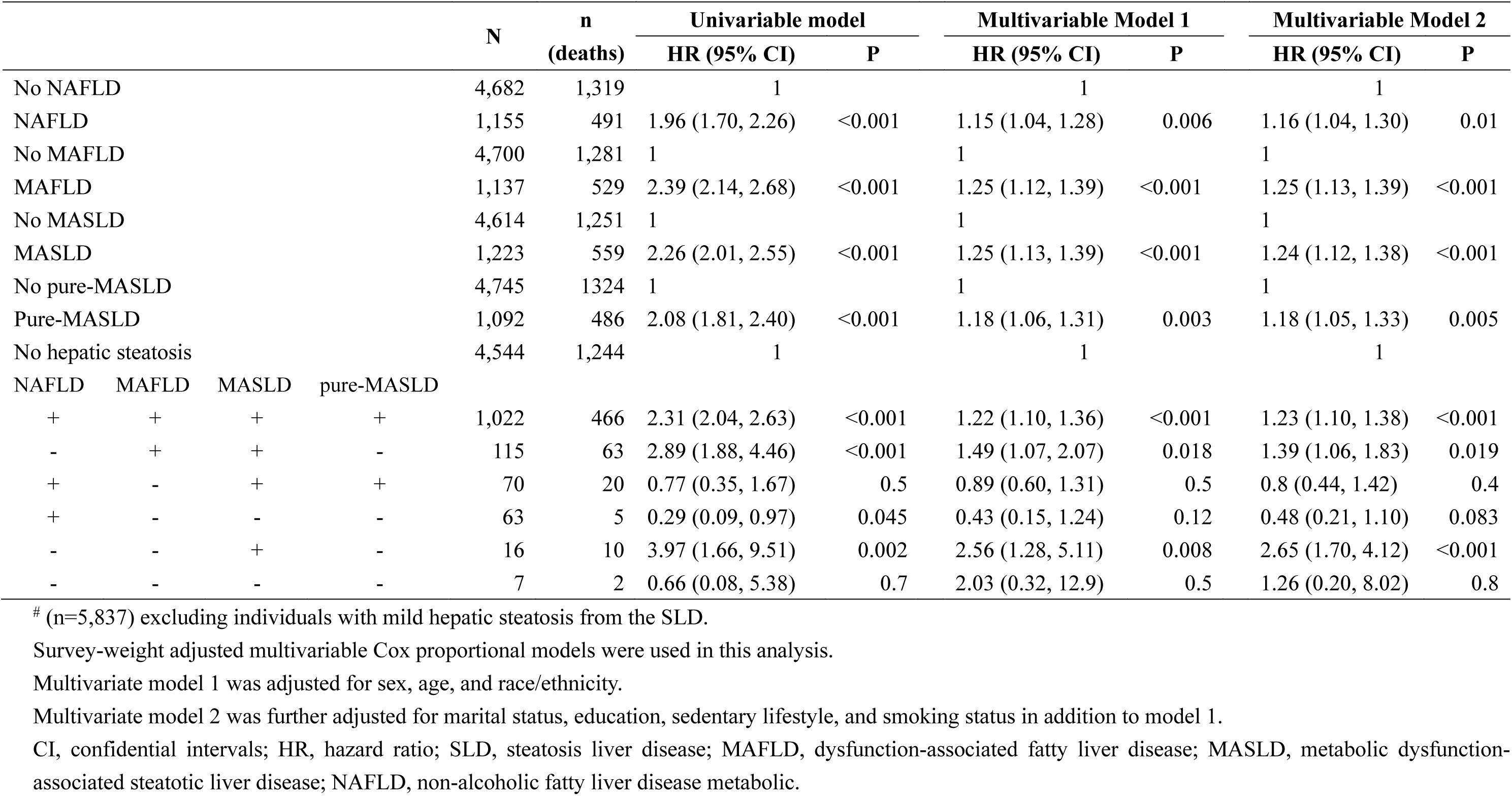
Association among NAFLD, MAFLD, MASLD and pure-MASLD status, and all-cause mortality in individuals with moderate-severe hepatic steatosis.

|  |  |  |  | N | n<br>(deaths) | Univariable model |  | Multivariable Model 1 |  | Multivariable Model 2 |  |
| --- | --- | --- | --- | --- | --- | --- | --- | --- | --- | --- | --- |
|  |  |  |  |  |  | HR (95% CI) | P | HR (95% CI) | P | HR (95% CI) | P |
| No NAFLD |  |  |  | 4,682 | 1,319 | 1 |  | 1 |  | 1 |  |
| NAFLD |  |  |  | 1,155 | 491 | 1.96 (1.70, 2.26) | <0.001 | 1.15 (1.04, 1.28) | 0.006 | 1.16 (1.04, 1.30) | 0.01 |
| No MAFLD |  |  |  | 4,700 | 1,281 | 1 |  | 1 |  | 1 |  |
| MAFLD |  |  |  | 1,137 | 529 | 2.39 (2.14, 2.68) | <0.001 | 1.25 (1.12, 1.39) | <0.001 | 1.25 (1.13, 1.39) | <0.001 |
| No MASLD |  |  |  | 4,614 | 1,251 | 1 |  | 1 |  | 1 |  |
| MASLD |  |  |  | 1,223 | 559 | 2.26 (2.01, 2.55) | <0.001 | 1.25 (1.13, 1.39) | <0.001 | 1.24 (1.12, 1.38) | <0.001 |
| No pure-MASLD |  |  |  | 4,745 | 1324 | 1 |  | 1 |  | 1 |  |
| Pure-MASLD |  |  |  | 1,092 | 486 | 2.08 (1.81, 2.40) | <0.001 | 1.18 (1.06, 1.31) | 0.003 | 1.18 (1.05, 1.33) | 0.005 |
| No hepatic steatosis |  |  |  | 4,544 | 1,244 | 1 |  | 1 |  | 1 |  |
| NAFLD | MAFLD | MASLD | pure-MASLD |  |  |  |  |  |  |  |  |
| + | + | + | + | 1,022 | 466 | 2.31 (2.04, 2.63) | <0.001 | 1.22 (1.10, 1.36) | <0.001 | 1.23 (1.10, 1.38) | <0.001 |
| - | + | + | - | 115 | 63 | 2.89 (1.88, 4.46) | <0.001 | 1.49 (1.07, 2.07) | 0.018 | 1.39 (1.06, 1.83) | 0.019 |
| + | - | + | + | 70 | 20 | 0.77 (0.35, 1.67) | 0.5 | 0.89 (0.60, 1.31) | 0.5 | 0.8 (0.44, 1.42) | 0.4 |
| + | - | - | - | 63 | 5 | 0.29 (0.09, 0.97) | 0.045 | 0.43 (0.15, 1.24) | 0.12 | 0.48 (0.21, 1.10) | 0.083 |
| - | - | + | - | 16 | 10 | 3.97 (1.66, 9.51) | 0.002 | 2.56 (1.28, 5.11) | 0.008 | 2.65 (1.70, 4.12) | <0.001 |
| - | - | - | - | 7 | 2 | 0.66 (0.08, 5.38) | 0.7 | 2.03 (0.32, 12.9) | 0.5 | 1.26 (0.20, 8.02) | 0.8 |

In addition, for the cause-specific mortality in different terms of SLD, similar results were also observed. Remarkably, we found a powerful protect effect to cardiovascular mortality in NAFLD+/MAFLD-/MASLD-/pure-MASLD- group (aHR: 0.02, 95% CI 0.00-0.18), but it was not occurred in cancer mortality analysis (Table S10). As for the advanced fibrosis, similar results were observed in intermediate and high NFS score groups (Table S11).

At last, we also conducted the subgroup analyses in this sensitivity analysis. Interestingly and importantly, all the NALFD, MAFLD, MASLD and pure-MASLD were associated with increased all-cause mortality in female, age 41-55 years, non-Hispanic white and never smoking subgroups (aHR: 1.19-1.61). Moreover, MAFLD and MASLD were associated with higher risk of all-cause mortality in BMI 18.5-24.9 kg/m^2^ and without diabetes subgroups (aHR: 1.18-1.52). (Figure 2B)

## Discussion

As the new term of MASLD was proposed to replace NAFLD and MAFLD recently, we evaluated the clinical characteristics and mortality outcomes about these different terms in this longitudinal, nationally representative, US population-based study. The main findings of our analysis were: 1) MASLD can identify more individuals with risk of all-cause mortality than MAFLD, due to broader definition of metabolic risk factors; 2) pure-MASLD and NAFLD showed high concordance in their clinical characteristics and outcomes, and both were not associated with all-cause mortality in total SLD individuals; 3) all the NAFLD, MAFLD, MASLD and pure-MASLD were associated with increased all-cause mortality in individuals with advanced fibrosis; 4) a higher risk of all-cause mortality of NAFLD, MAFLD, MASLD and pure-MASLD were also observed in female, age 41-55 years, non-Hispanic white, and never smoking subgroups when focused on moderate-severe hepatic steatosis.

Regarding to the relatively strict and complex of MAFLD definition ^16^, MASLD simplified the number of metabolic risk factors, and relaxed the range of some criteria, such as the waist circumference ^2^. Hence, MASLD will include more SLD individuals than MAFLD in theory, and this was proved by our result, that is, MASLD can identify about 3% more individuals than MAFLD (29.04% vs. 26.13%). This phenomenon was also observed in a previous study from Hong Kong cohort (MASLD: 26.7% vs. MAFLD: 25.9%) ^5^. Although several previous studies had assessed the association between MAFLD and all-cause mortality based on NHAENS III data ^19 21–23^, the conclusions are still controversial, due to different adjusted models chose. Considering the possible pitfalls in the adjustment model choosing ^20^, we just adjusted the sociodemographic characteristics and lifestyle risk factors of individuals, and any variable used in their definitions were discarded. In this scenario, our result showed a significant higher risk of all-cause mortality in individuals with MAFLD, which was similar with a recent study using the same strategy ^11^.

Importantly, we found that MASLD was also significantly associated with increased all-cause mortality after adjustment. This result indicated that new term of MASLD is better identify people at risk of all-cause mortality than MAFLD. Because no study focused on the clinical outcome of MASLD in a broad sense before, more studies are needed to verify the result.

To better distinguish the effect of alcohol intake from MASLD, the Delphi consensus further classified MASLD into pure-MASLD, MetALD, and MASLD with other etiologies. According to the definition, individuals with excessive alcohol consumption were excluded from both pure-MASLD and NAFLD, but pure-MASLD was restricted by the metabolic risk factors definition ^2 15^. In this study, there were 10.1% individuals with NAFLD cannot be classified into pure-MASLD. This was different with previous studies, which showed extremely high (over 99%) accordance between NAFLD and pure-MASLD in cohorts from France ^6^ or Sweden ^8^. This discrepancy may be attributed to the differences of region, race, and enrollment time of participants. As expected, all the individuals with pure-MASLD can be classified into NAFLD. For the mortality outcomes, previous NHANES III based studies showed NAFLD per se was not associated with increased risk of all-cause and cause-specific mortality, which were in line with our result with a more extended follow-up period of the median 27 years ^19 21 24^. Similarly, we found that pure-MASLD was also not associated with all-cause mortality after adjustment, which was in accordance with a recent study ^10^. But Zhao et al. reported a higher risk of all-cause mortality in individuals with pure-MASLD ^11^. This may due to more participants without alcohol intake information were included in their study, which will result in some people who have no information about alcohol consumption but indeed have excessive alcohol consumption being classified into pure-MASLD group. Our results suggested that NAFLD and pure-MASLD were highly similar in characteristics and clinical outcomes ^9^, hence, old NAFLD data could still be used and new biomarkers to differentiate NAFLD and pure-MASLD may be not needed.

Next, we evaluated the mortality among the different combinations of NAFLD, MAFLD, MASLD and pure-MASLD. Due to other confounders of SLD, such as viral hepatitis and low BMI, has been excluded in the main analysis, we can better to focus on the effect of metabolic risk factors and alcohol intake on the outcomes of the four terms. Both NAFLD+/MAFLD+/MASLD+/pure- MASLD+ and NAFLD-/MAFLD+/MASLD+/pure-MASLD- were associated with increased all-cause mortality, and the later showed a larger effect. This indicated that metabolic dysregulation met the criteria of MAFLD alone will increase the mortality risk, and excessive alcohol intake plus metabolic dysregulation will further strengthen the risk of mortality, which was also observed in previous study ^19^. Interestingly, we found that NAFLD+/MAFLD-/MASLD+/pure-MASLD+, but not NAFLD+/MAFLD-/MASLD-/pure-MASLD- was associated with lower risk of all-cause and cause-specific mortality. This indicated that instead of no metabolic risk, only slight metabolic risk (MAFLD-/MASLD+) was a protected effect to mortality in individuals with SLD. It is hard for us to explain this odd result now, may be related to the small number of each group, and studies with larger sample size are needed to clarify this. In addition, both NAFLD-/MAFLD-/MASLD+/pure-MASLD- and NAFLD-/MAFLD-/MASLD-/pure-MASLD- were not associated with all-cause and cause-specific mortality. This suggesting that excessive alcohol consumption alone or plus slight metabolic risk will not increase the risk of mortality. Taken together, the increased risk of mortality in SLD was only related to the extent of metabolic dysregulation, and alcohol intake plays a boost role in this.

Except for metabolism and alcohol, fibrosis is another factor related to poor progress ^25^. Here, we used the NFS score, as a surrogate index due to lack of the liver biopsy, to assess the possibility of advanced fibrosis ^17^. Previous study showed that advanced fibrosis in MAFLD, but not NAFLD, was associated with a higher risk of all-cause mortality ^19^. In our analysis, not only MAFLD, but also NAFLD, MASLD and pure-MASLD were associated with increased all-cause mortality, and the effects were larger with higher NFS score. Hence, even in the new term of MASLD, individuals with advanced fibrosis are still a special population needed to pay more attention.

In subgroup analyses, we noticed that both MAFLD and MASLD were associated with increased all-cause mortality in middle age (41-55 years) and non-Hispanic white subgroups. And both NAFLD and pure-MASLD were not associated with all-cause mortality in any subgroup, which was similar with a previous study on NAFLD ^24^. Notably, more powerful effects of all-cause-mortality risk were observed among the four terms after excluding individuals with mild hepatic steatosis from those with SLD. In this sensitivity analysis, individuals with NAFLD, MAFLD, MASLD and pure-MASLD had higher risk of all-cause-mortality in female, middle age, non-Hispanic white and never smoking subgroups. The reasons for these phenomena are complex and unclear. For the age, we speculated the reason may be that the younger subgroup had a lower mortality and the older subgroup had a higher mortality, which may counteract the effects of NAFLD, MAFLD, MASLD and pure-MASLD. Similarly, we guessed that individuals with the history of smoking (irrespective of previous or current smoking) had a higher mortality than never smoking subgroup ^26^, which may weaken the discrepancy caused by NAFLD, MAFLD, MASLD and pure-MASLD. But for sex and race, it is hard to confirm the specific cause now. The results in subgroup analyses needed more studies to investigate and verify in future. All in all, subgroup and sensitivity analyses showed the increased all-cause mortality among NAFLD, MAFLD, MASLD and pure-MASLD were related to sex, age, race, smoking status and the extent of hepatic steatosis.

Our study has some strengths. First, we comprehensively compared the clinical characteristics and outcomes of NAFLD, MAFLD, MASLD and pure-MASLD using the NHANES III data, a large nationally representative sample of the US population with a long follow-up period for mortality (median of 27 years). Second, we conducted a strict screen strategy excluding individuals without alcohol consumption information, or with other confounders (such as pregnancy, abnormal liver enzymes levels, viral hepatitis, or BMI less than 18.5 kg/m^2^), so that we can focus on the effect of metabolic risk factors and alcohol intake on the mortality of these terms. Third, we first found that the increased all-cause mortality among NAFLD, MAFLD, MASLD and pure-MASLD were associated with sex, age, race, smoking status and the extent of hepatic steatosis in subgroup and sensitivity analyses. Although the specific cause is unclear, more attention and earlier intervention should be paid to these special populations in future.

Meanwhile, our study also has some limitations. First, due to the participants in NHANES III were included between 1988-1994, the prevalence of NAFLD, MAFLD, MASLD and pure-MASLD may not reflect the current extent. Second, due to the cross-sectional design of NHANES, we were unable to assess dynamic changes in NAFLD, MAFLD, MASLD and pure-MASLD status over time. Third, individuals with viral hepatitis or low BMI were added into the entire population in sensitivity analysis, and similar results were observed. But considering the number of these individuals were relatively small, the results should be interpreted with caution. Hence, the specific effect of viral hepatitis on the mortality outcomes in SLD is still need to be verified in studies with larger samples, especially in Asia and Africa where the prevalence of viral hepatitis was high ^27^. Despite some of these limitations, we believe our findings are still important and meaningful, benefited of a large population-based study using a nationally representative sample and sufficient follow-up period.

In conclusion, the definition of MASLD was better than MAFLD to identify more individuals with poor progress, whereas, pure-MASLD had similar characteristics and mortality outcomes with NAFLD. The increased all-cause mortality of SLD was only associated with the extent of metabolic dysfunction, but excessive alcohol intake will enlarge this effect. Moreover, higher risk of all-cause mortality of NAFLD, MAFLD, MASLD and pure-MASLD were also observed in individuals with advanced fibrosis and moderate-severe of hepatic steatosis, and in female, middle age, non-Hispanic white, and never smoking subgroups. Well-designed prospective studies with larger samples are needed to verify our observations in future.

## Data Availability

All data produced in the present work are contained in the manuscript.

## Acknowledgement

We gratefully acknowledge the contribution of the participants of the NHANES cohort, research assistants, and facilitating personnel. And this work was supported by the first batch of key disciplines on public health in Chongqing, Health Commission of Chongqing, China.

## Author contributions

Rui Song, Zhao Li and Yingzhi Zhang was involved in acquisition of data, data analysis and interpretation, drafting of the manuscript, and study supervision. Jiahe Tan were involved in interpretation of data and critical revision of the manuscript. Zhiwei Chen was involved in study concept and design, interpretation of data, drafting of the manuscript, critical revision of the manuscript, and study supervision. All authors contributed to the article and approved the submitted version.

