## Supplementary Materials for "Comparison of NAFLD, MAFLD, MASLD and pure-MASLD characteristics and mortality outcomes in United States adults"

**Supplementary methods**

**Other definitions**

Hypertension was defined as systolic blood pressure ≥140 mm Hg or diastolic blood pressure ≥90 mm Hg, and/or self-reported doctor diagnosis, and/or antihypertensive drug use [1].

Diabetes was defined as fasting glucose ≥126 mg/dl, hemoglobin A1c ≥6.5%, and/or self-reported doctor diagnosis, and/or treatment with anti-diabetic medication [2].

Smoking status was categorized as never, past, and current smoker. Never smokers replied no to the question: "Have you smoked at least 100 cigarettes during your entire life?" Current smokers were defined as those who reported ongoing smoking among individuals who had smoked at least 100 cigarettes in their lifetime [2].

Alcohol consumption was calculated as "gram of alcohol intake per day (g/d)". In detail, average daily number of alcoholic drinks was estimated by multiplying the number of drinking days over the past 12 months and the number of drinks, on average, on a drinking day and dividing by 365 [1]. According to US standards, alcoholic drinks counted as 14 g of alcohol each [3]. In addition, never drinkers replied no to the question: "In your entire life, have you had at least 12 drinks of any kind of alcoholic beverage?" Average daily alcohol consumption was defined as zero in never drinkers in this study. Excessive alcohol consumption was defined as ≥20 g/d for females and ≥30 g/d for males [4].

Sedentary behavior was defined as if individuals answered 'no' to all questions about engaging in any of the following physical activities over the last month: jog or run, cycle, swim, aerobics, other dancing, calisthenics, garden or yard work, weight lifting, or other sports [1].

HBV was defined as hepatitis B surface antigen-positive. HCV was defined as hepatitis C antibody-positive.

MAFLD was defined as the presence of SLD with metabolic dysfunction, which comprises either overweight or obese (BMI ≥25 kg/m$^2$), diabetes mellitus, or a combination of at least 2 of following metabolic risk abnormalities: (1) waist circumference ≥102 cm for males and ≥88 cm for females, (2) blood pressure ≥130/85 mmHg or antihypertensive drug treatment, (3) plasma triglycerides ≥150 mg/dl or lipid-lowering drug treatment, (4) plasma HDL-cholesterol <40 mg/dl for men and <50 mg/dl for women or lipid-lowering drug treatment, (5) prediabetes defined as fasting glucose 100-125 mg/dl or hemoglobin A1c 5.7%-6.4%, (6) homeostasis model assessment of insulin resistance score ≥2.5, (7) C-reactive protein (CRP) level >2 mg/L [5].

MASLD was defined as the presence of SLD combined with at least 1 of the 5 following cardiometabolic adult criteria: (1) BMI ≥25 kg/m$^2$ or waist circumference ≥94 cm for males and ≥80 cm for females, (2) fasting glucose ≥100 mg/dl or 2-hour post-load glucose levels ≥140mg/dl or hemoglobin A1c ≥5.7% or diabetes mellitus or treatment for diabetes mellitus, (3) blood pressure ≥130/85 mmHg or antihypertensive drug treatment, (4) fasting plasma triglycerides ≥150 mg/dl or lipid-lowering treatment, (5) plasma HDL-cholesterol <40 mg/dl for men and <50 mg/dl for women or lipid-lowering treatment [4].

Alcoholic liver disease (ALD) was defined as the presence of SLD and reported higher alcohol consumption (>50 g/d for females and >60 g/d for males) in this study [6].

In NHANES III, mortality was evaluated by Underlying Cause of Death 113 (UCOD_113) code. All-cause mortality was classified based on the ICD-9 for deaths through 1998, and ICD-10 for deaths from 1999 to 2019. Cause-specific mortalities were assessed as cardiovascular disease (UCOD_113 code: 55-64, 70) and cancer (UCOD_113 code: 19-43) [2].

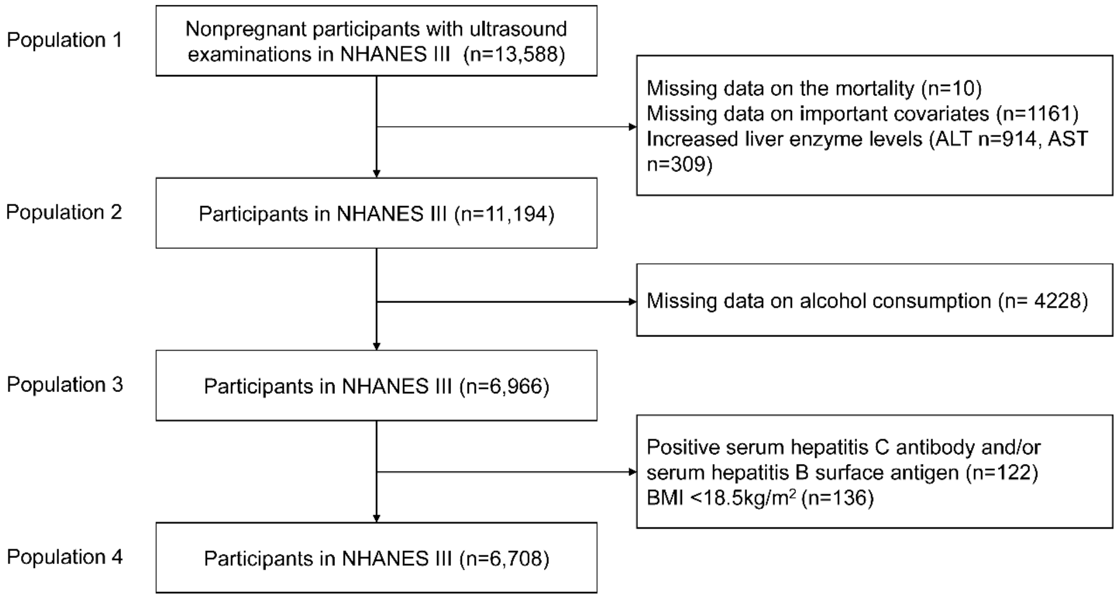

**Figure S1. Flow diagram of participants through the study.**

**Table S1. Comparison of demographic and clinical characteristics among population stepwise screened in the study.**

| Characteristics | Population 1# (n=13,588) | Population 2# (n=11,194) | Population 3# (n=6,966) | Population 4# (n=6,708) |
|---|---|---|---|---|
| Age (years) | 40.0 (30.0, 54.0) | 40.0 (30.0, 54.0) | 38.0 (29.0, 51.0) | 38.0 (29.0, 52.0) |
| Sex (male) | 6,475 (49%) | 5,252 (49%) | 3,517 (52%) | 3,390 (53%) |
| Race/ethnicity | | | | |
| Non-Hispanic white | 4,972 (76%) | 4,348 (77%) | 2,758 (77%) | 2,663 (78%) |
| Non-Hispanic black | 4,042 (5.4%) | 3,159 (4.9%) | 1,965 (4.7%) | 1,923 (4.8%) |
| Mexican-American | 3,990 (11%) | 3,217 (10%) | 1,928 (9.4%) | 1,826 (9.3%) |
| Others | 574 (8.1%) | 470 (7.7%) | 315 (8.4%) | 296 (8.2%) |
| Body mass index (kg/m$^2$) | 25.5 (22.6, 29.5) | 25.4 (22.5, 29.1) | 25.0 (22.3, 28.4) | 25.2 (22.6, 28.6) |
| Waist circumference (cm) | 91 (81, 101) | 90 (80, 100) | 89 (80, 99) | 90 (80, 99) |
| Triglycerides (mg/dL) | 1.2 (0.9, 1.9) | 1.2 (0.9, 1.8) | 1.2 (0.8, 1.7) | 1.2 (0.9, 1.8) |
| HDL-cholesterol (mg/dL) | 1.2 (1.0, 1.5) | 1.2 (1.0, 1.5) | 1.3 (1.1, 1.6) | 1.3 (1.1, 1.6) |
| C-reactive protein (mg/dL) | 0.2 (0.2, 0.3) | 0.2 (0.2, 0.3) | 0.2 (0.2, 0.2) | 0.2 (0.2, 0.2) |
| Glucose (mol/L) | 5.1 (4.8, 5.5) | 5.1 (4.8, 5.5) | 5.1 (4.8, 5.5) | 5.1 (4.8, 5.5) |
| Insulin (uU/mL) | 8.2 (5.8, 12.1) | 7.9 (5.7, 11.5) | 7.7 (5.6, 10.9) | 7.7 (5.6, 11.0) |
| Hemoglobin A1c (%) | 5.2 (4.9, 5.5) | 5.2 (4.9, 5.5) | 5.2 (4.9, 5.5) | 5.2 (4.9, 5.5) |
| HOMA_IR | 1.9 (1.3, 2.9) | 1.8 (1.3, 2.8) | 1.7 (1.2, 2.6) | 1.7 (1.2, 2.6) |
| Hepatic steatosis (%) | 4,947 (34%) | 3,794 (32%) | 2,241 (30%) | 2,164 (30%) |

Data are shown as the median (interquartile range) or unweighted frequency counts (weighted percentage) as appropriate.

### Population 1, nonpregnant participants with available ultrasound examination results in NHANES III; Population 2, excluding participants with missing data or increased liver enzyme levels; Population 3, further excluding participants without alcohol consumption information; Population 4, further excluding participants with viral hepatitis or low body mass index.

HDL, high-density lipoprotein; HOMA-IR, homeostasis model assessment of insulin resistance.

**Table S2. Baseline characteristics of NAFLD, MAFLD, MASLD and pure-MASLD.**

| Characteristics | No hepatic steatosis (n=4,544) | SLD (n=2,164) | P | NAFLD (n=1,937) | MAFLD (n=1,735) | MASLD (n=1,948) | Pure-MASLD (n=1,741) |
|---|---|---|---|---|---|---|---|
| Age (years) | 36.0 (28.0, 48.0) | 42.0 (32.0, 57.0) | <0.001 | 42.0 (32.0, 57.0) | 47.0 (36.0, 60.0) | 45.0 (34.0, 58.0) | 45.0 (34.0, 58.0) |
| Sex (male) | 2,301 (52%) | 1,089 (54%) | | 916 (51%) | 908 (59%) | 988 (56%) | 830 (53%) |
| Race/ethnicity | | | 0.11 | | | | |
| Non-Hispanic white | 1,814 (78%) | 849 (78%) | | 752 (77%) | 673 (76%) | 760 (77%) | 672 (76%) |
| Non-Hispanic black | 1,198 (4.5%) | 725 (5.6%) | | 653 (5.8%) | 622 (6.1%) | 673 (5.8%) | 603 (5.9%) |
| Mexican-American | 1,324 (9.8%) | 502 (8.1%) | | 449 (8.2%) | 384 (8.0%) | 433 (8.1%) | 388 (8.2%) |
| Others | 208 (8.1%) | 88 (8.4%) | | 83 (9.0%) | 74 (9.4%) | 82 (8.9%) | 78 (9.7%) |
| Body mass index (kg/m$^2$) | 24.6 (22.3, 27.3) | 27.3 (24.2, 31.2) | <0.001 | 27.5 (24.2, 31.2) | 29.3 (26.4, 32.4) | 28.3 (25.4, 31.9) | 28.4 (25.5, 31.9) |
| Waist circumference (cm) | 88 (79, 96) | 97 (85, 106) | <0.001 | 97 (85, 106) | 101 (94, 110) | 99 (90, 108) | 99 (90, 108) |
| ≥12 years education | 3,094 (82%) | 1,259 (75%) | <0.001 | 1,123 (75%) | 980 (74%) | 1,109 (75%) | 985 (74%) |
| Marital status | 2,798 (65%) | 1,459 (72%) | 0.001 | 1,315 (73%) | 1,214 (74%) | 1,332 (72%) | 1,198 (73%) |
| Hypertension | 1,140 (20%) | 834 (37%) | <0.001 | 718 (34%) | 796 (46%) | 827 (41%) | 713 (39%) |
| Diabetes | 219 (2.8%) | 324 (9.6%) | <0.001 | 299 (10.0%) | 323 (13%) | 324 (11%) | 299 (11%) |
| Smoking status | | | 0.010 | | | | |
| Never | 2,297 (46%) | 1,080 (47%) | | 1,028 (49%) | 867 (46%) | 964 (46%) | 917 (48%) |
| Past smoker | 866 (22%) | 545 (27%) | | 473 (26%) | 491 (31%) | 521 (29%) | 451 (28%) |
| Current smoker | 1,381 (32%) | 539 (27%) | | 436 (25%) | 395 (24%) | 463 (25%) | 373 (24%) |
| Alcohol consumption (g/d) | 4 (1, 12) | 4 (0, 12) | 0.072 | 2 (0, 8) | 4 (0, 12) | 4 (0, 12) | 2 (0, 8) |
| Sedentary lifestyle | 1,061 (16%) | 655 (21%) | <0.001 | 598 (21%) | 562 (23%) | 612 (22%) | 561 (23%) |
| Triglycerides (mg/dL) | 1.1 (0.8, 1.6) | 1.5 (1.0, 2.3) | <0.001 | 1.5 (1.0, 2.3) | 1.7 (1.2, 2.5) | 1.6 (1.1, 2.4) | 1.6 (1.1, 2.4) |
| HDL-cholesterol (mg/dL) | 1.3 (1.1, 1.6) | 1.2 (1.0, 1.5) | <0.001 | 1.2 (1.0, 1.4) | 1.1 (0.9, 1.4) | 1.2 (1.0, 1.4) | 1.1 (0.9, 1.4) |
| C-reactive protein (mg/dL) | 0.2 (0.2, 0.2) | 0.2 (0.2, 0.3) | <0.001 | 0.2 (0.2, 0.4) | 0.2 (0.2, 0.4) | 0.2 (0.2, 0.4) | 0.2 (0.2, 0.4) |
| Glucose (mol/L) | 5.1 (4.8, 5.4) | 5.2 (4.9, 5.6) | <0.001 | 5.2 (4.9, 5.7) | 5.3 (5.0, 5.8) | 5.3 (5.0, 5.7) | 5.3 (5.0, 5.8) |

| | | | | | | | |
|---|---|---|---|---|---|---|---|
| Insulin (uU/mL) | 7.0 (5.4, 9.7) | 9.5 (7.0, 13.9) | <0.001 | 10 (7, 14) | 11 (8, 15) | 10 (8, 15) | 11 (8, 15) |
| Hemoglobin A1c (%) | 5.1 (4.8, 5.4) | 5.2 (4.9, 5.6) | <0.001 | 5.3 (4.9, 5.6) | 5.3 (5.0, 5.7) | 5.3 (5.0, 5.7) | 5.3 (5.0, 5.7) |
| HOMA-IR | 1.6 (1.2, 2.3) | 2.3 (1.6, 3.5) | <0.001 | 2.4 (1.6, 3.6) | 2.7 (1.9, 4.0) | 2.5 (1.7, 3.7) | 2.5 (1.8, 3.8) |

Data are shown as the median (interquartile range) or unweighted frequency counts (weighted percentage) as appropriate. The Wilcoxon test for continuous variables and the Chi-square test for categorical variables were used in this analysis.

HDL, high-density lipoprotein; HOMA-IR, homeostasis model assessment of insulin resistance; SLD, steatosis liver disease; MAFLD, dysfunction-associated fatty liver disease; MASLD, metabolic dysfunction-associated steatotic liver disease; NAFLD, non-alcoholic fatty liver disease metabolic.

**Table S3. Association different subgroups of SLD and all-cause mortality according to the definition of MASLD.**

| | N | n | Univariable model | | Multivariable Model 1 | | Multivariable Model 2 | |
|---|---|---|---|---|---|---|---|---|
| | | (deaths) | HR (95% CI) | P | HR (95% CI) | P | HR (95% CI) | P |
| No hepatic steatosis | 4,544 | 1,244 | 1 | | 1 | | 1 | |
| Pure_MASLD | 1,741 | 693 | 1.75 (1.55-1.98) | <0.001 | 1.1 (0.98-1.23) | 0.11 | 1.09 (0.98-1.22) | 0.13 |
| MetALD | 157 | 70 | 2.18 (1.55-3.06) | <0.001 | 1.29 (0.88-1.88) | 0.2 | 1.17 (0.84-1.64) | 0.3 |
| MASLD_ALD | 50 | 29 | 2.39 (0.86-6.60) | 0.094 | 2.14 (1.22-3.76) | 0.008 | 2.10 (1.22-3.60) | 0.007 |
| Steatosis without MASLD | 216 | 25 | 0.51 (0.30-0.86) | 0.012 | 1.12 (0.71-1.78) | 0.6 | 1.18 (0.75-1.85) | 0.5 |

Survey-weight adjusted multivariable Cox proportional models were used in this analysis.

Multivariate model 1 was adjusted for sex, age, and race/ethnicity.

Multivariate model 2 was further adjusted for marital status, education, sedentary lifestyle, and smoking status in addition to model 1.

ALD, alcohol-related liver disease; CI, confidential intervals; HR, hazard ratio; SLD, steatosis liver disease; MASLD, metabolic dysfunction-associated steatotic liver disease.

**Table S4. Association among NAFLD, MAFLD, MASLD and pure-MASLD status, and all-cause mortality (for the sensitivity analysis 1[#]).**

| | | | | N | n (deaths) | Univariable model | | Multivariable Model 1 | | Multivariable Model 2 | |
|---|---|---|---|---|---|---|---|---|---|---|---|
| | | | | | | HR (95% CI) | P | HR (95% CI) | P | HR (95% CI) | P |
| No NAFLD | | | | 4,993 | 1,424 | 1 | | 1 | | 1 | |
| NAFLD | | | | 1,973 | 719 | 1.47 (1.30, 1.67) | <0.001 | 1.05 (0.93, 1.18) | 0.4 | 1.06 (0.94, 1.18) | 0.3 |
| No MAFLD | | | | 5,187 | 1,379 | 1 | | 1 | | 1 | |
| MAFLD | | | | 1,779 | 764 | 2.08 (1.85, 2.34) | <0.001 | 1.18 (1.05, 1.32) | 0.004 | 1.2 (1.08, 1.34) | <0.001 |
| No MASLD | | | | 4,965 | 1,331 | 1 | | 1 | | 1 | |
| MASLD | | | | 2,001 | 812 | 1.83 (1.65, 2.02) | <0.001 | 1.12 (1.01, 1.25) | 0.031 | 1.11 (1.00, 1.22) | 0.041 |
| No pure-MASLD | | | | 5,179 | 1,434 | 1 | | 1 | | 1 | |
| Pure-MASLD | | | | 1,787 | 709 | 1.69 (1.49, 1.93) | <0.001 | 1.06 (0.95, 1.19) | 0.3 | 1.06 (0.95, 1.18) | 0.3 |
| No hepatic steatosis | | | | 4,725 | 1,303 | 1 | | 1 | | 1 | |
| NAFLD | MAFLD | MASLD | pure-MASLD | | | | | | | | |
| + | + | + | + | 1,580 | 668 | 1.96 (1.72, 2.24) | <0.001 | 1.13 (1.01, 1.27) | 0.032 | 1.15 (1.04, 1.29) | 0.01 |
| - | + | + | - | 199 | 96 | 2.13 (1.49, 3.06) | <0.001 | 1.45 (1.05, 1.99) | 0.022 | 1.38 (1.06, 1.79) | 0.017 |
| + | - | + | + | 183 | 31 | 0.5 (0.33, 0.77) | 0.001 | 0.6 (0.43, 0.85) | 0.004 | 0.48 (0.30, 0.77) | 0.003 |
| + | - | - | - | 210 | 20 | 0.46 (0.25, 0.83) | 0.011 | 0.95 (0.59, 1.52) | 0.8 | 1.05 (0.67, 1.64) | 0.8 |
| - | - | + | - | 39 | 17 | 1.68 (0.74, 3.80) | 0.2 | 1.43 (0.69, 2.95) | 0.3 | 1.04 (0.40, 2.70) | >0.9 |
| - | - | - | - | 30 | 8 | 0.69 (0.24, 1.99) | 0.5 | 2.32 (0.86, 6.24) | 0.1 | 1.51 (0.56, 4.10) | 0.4 |

[#] (n=6,966) including individuals with viral hepatitis or low BMI (<18.5 kg/m$^2$).

Survey-weight adjusted multivariable Cox proportional models were used in this analysis.

Multivariate model 1 was adjusted for sex, age, and race/ethnicity.

Multivariate model 2 was further adjusted for marital status, education, sedentary lifestyle, and smoking status in addition to model 1.

CI, confidential intervals; HR, hazard ratio; MAFLD, dysfunction-associated fatty liver disease; MASLD, metabolic dysfunction-associated steatotic liver disease; NAFLD, non-alcoholic fatty liver disease metabolic.

**Table S5. Association different subgroups of SLD and all-cause mortality according to the definition of MASLD (for the sensitivity analysis 1#).**

| | N | n (deaths) | Univariable model | | Multivariable Model 1 | | Multivariable Model 2 | |
|---|---|---|---|---|---|---|---|---|
| | | | HR (95% CI) | P | HR (95% CI) | P | HR (95% CI) | P |
| No hepatic steatosis | 4,725 | 1,303 | 1 | | 1 | | 1 | |
| Pure_MASLD | 1,787 | 709 | 1.71 (1.52, 1.94) | <0.001 | 1.09 (0.97, 1.22) | 0.13 | 1.08 (0.97, 1.21) | 0.14 |
| MetALD | 160 | 72 | 2.24 (1.58, 3.18) | <0.001 | 1.31 (0.89, 1.93) | 0.2 | 1.2 (0.85, 1.69) | 0.3 |
| MASLD_ALD | 54 | 31 | 2.25 (0.86, 5.90) | 0.1 | 2.11 (1.22, 3.65) | 0.008 | 2.07 (1.23, 3.50) | 0.006 |
| Steatosis without MASLD | 240 | 28 | 0.49 (0.29, 0.82) | 0.007 | 1.08 (0.70, 1.68) | 0.7 | 1.12 (0.73, 1.72) | 0.6 |

### (n=6,966) including individuals with viral hepatitis or low BMI (<18.5 kg/$m^2$).

Survey-weight adjusted multivariable Cox proportional models were used in this analysis.

Multivariate model 1 was adjusted for sex, age, and race/ethnicity.

Multivariate model 2 was further adjusted for marital status, education, sedentary lifestyle, and smoking status in addition to model 1.

ALD, alcohol-related liver disease; CI, confidential intervals; HR, hazard ratio; SLD, steatosis liver disease; MASLD, metabolic dysfunction-associated steatotic liver disease.

**Table S6. Association among NAFLD, MAFLD, MASLD and pure-MASLD status, and cardiovascular disease and cancer-related mortality (for the sensitivity analysis 1[#]).**

| | | | | N | n (deaths) | Univariable model | | Multivariable Model 1 | | Multivariable Model 2 | |
|---|---|---|---|---|---|---|---|---|---|---|---|
| | | | | | | HR (95% CI) | P | HR (95% CI) | P | HR (95% CI) | P |
| **Cardiovascular mortality** | | | | | | | | | | | |
| No NAFLD | | | | 4,993 | 356 | 1 | | 1 | | 1 | |
| NAFLD | | | | 1,973 | 193 | 1.44 (1.08, 1.92) | 0.014 | 0.97 (0.73, 1.29) | 0.8 | 0.97 (0.72, 1.30) | 0.8 |
| No MAFLD | | | | 5,187 | 341 | 1 | | 1 | | 1 | |
| MAFLD | | | | 1,779 | 208 | 2.25 (1.73, 2.93) | <0.001 | 1.19 (0.93, 1.52) | 0.2 | 1.22 (0.95, 1.57) | 0.12 |
| No MASLD | | | | 4,965 | 332 | 1 | | 1 | | 1 | |
| MASLD | | | | 2,001 | 217 | 1.88 (1.43, 2.47) | <0.001 | 1.09 (0.84, 1.40) | 0.5 | 1.07 (0.81, 1.40) | 0.7 |
| No pure-MASLD | | | | 5,179 | 359 | 1 | | 1 | | 1 | |
| Pure-MASLD | | | | 1,787 | 190 | 1.72 (1.30, 2.28) | <0.001 | 1.02 (0.77, 1.35) | >0.9 | 1 (0.74, 1.35) | >0.9 |
| No hepatic steatosis | | | | 4,725 | 327 | 1 | | 1 | | 1 | |
| NAFLD | MAFLD | MASLD | pure-MASLD | | | | | | | | |
| + | + | + | + | 1,580 | 185 | 2.05 (1.56, 2.71) | <0.001 | 1.11 (0.84, 1.46) | 0.5 | 1.13 (0.85, 1.49) | 0.4 |
| - | + | + | - | 199 | 23 | 2.25 (1.21, 4.19) | 0.011 | 1.49 (0.76, 2.92) | 0.2 | 1.42 (0.76, 2.65) | 0.3 |
| + | - | + | + | 183 | 5 | 0.18 (0.04, 0.85) | 0.03 | 0.22 (0.05, 0.99) | 0.048 | 0.16 (0.03, 0.80) | 0.026 |
| + | - | - | - | 210 | 3 | 0.11 (0.02, 0.63) | 0.014 | 0.25 (0.04, 1.58) | 0.14 | 0.29 (0.05, 1.60) | 0.2 |
| - | - | + | - | 39 | 4 | 1.07 (0.29, 3.94) | >0.9 | 0.82 (0.18, 3.69) | 0.8 | 0.72 (0.18, 2.92) | 0.6 |
| - | - | - | - | 30 | 2 | 1 (0.16, 6.41) | >0.9 | 4.74 (0.83, 26.9) | 0.079 | 3.02 (0.53, 17.1) | 0.2 |
| **Cancer mortality** | | | | | | | | | | | |
| No NAFLD | | | | 4,993 | 384 | 1 | | 1 | | 1 | |
| NAFLD | | | | 1,973 | 167 | 1.25 (0.92, 1.70) | 0.2 | 0.93 (0.68, 1.26) | 0.6 | 0.98 (0.72, 1.33) | >0.9 |

| | | | | | | | | | | | |
|---|---|---|---|---|---|---|---|---|---|---|---|
| No MAFLD | | | | 5,187 | 372 | 1 | | 1 | | 1 | |
| MAFLD | | | | 1,779 | 179 | 1.84 (1.42, 2.38) | <0.001 | 1.1 (0.85, 1.43) | 0.5 | 1.19 (0.91, 1.57) | 0.2 |
| No MASLD | | | | 4,965 | 358 | 1 | | 1 | | 1 | |
| MASLD | | | | 2,001 | 193 | 1.6 (1.28, 2.00) | <0.001 | 1.03 (0.81, 1.32) | 0.8 | 1.07 (0.84, 1.36) | 0.6 |
| No pure-MASLD | | | | 5,179 | 384 | 1 | | 1 | | 1 | |
| Pure-MASLD | | | | 1,787 | 167 | 1.42 (1.07, 1.89) | 0.015 | 0.94 (0.71, 1.25) | 0.7 | 0.99 (0.75, 1.31) | >0.9 |
| No hepatic steatosis | | | | 4,725 | 350 | 1 | | 1 | | 1 | |
| NAFLD | MAFLD | MASLD | pure-MASLD | | | | | | | | |
| + | + | + | + | 1,580 | 152 | 1.67 (1.22, 2.28) | 0.002 | 1.02 (0.75, 1.38) | >0.9 | 1.1 (0.81, 1.50) | 0.5 |
| - | + | + | - | 199 | 27 | 2.38 (1.37, 4.13) | 0.002 | 1.61 (0.95, 2.71) | 0.077 | 1.6 (0.94, 2.71) | 0.081 |
| + | - | + | + | 183 | 10 | 0.4 (0.16, 0.98) | 0.044 | 0.48 (0.20, 1.12) | 0.091 | 0.37 (0.14, 0.98) | 0.045 |
| + | - | - | - | 210 | 5 | 0.48 (0.14, 1.61) | 0.2 | 0.89 (0.27, 2.95) | 0.9 | 0.98 (0.29, 3.35) | >0.9 |
| - | - | + | - | 39 | 4 | 1.43 (0.39, 5.17) | 0.6 | 1.23 (0.29, 5.18) | 0.8 | 0.8 (0.18, 3.62) | 0.8 |
| - | - | - | - | 30 | 3 | 1.34 (0.34, 5.31) | 0.7 | 3.16 (0.82, 12.2) | 0.1 | 2.01 (0.53, 7.68) | 0.3 |

### (n=6,966) including individuals with viral hepatitis or low BMI (<18.5 kg/$m^2$).

Survey-weight adjusted multivariable Cox proportional models were used in this analysis.

Multivariate model 1 was adjusted for sex, age, and race/ethnicity.

Multivariate model 2 was further adjusted for marital status, education, sedentary lifestyle, and smoking status in addition to model 1.

CI, confidential intervals; HR, hazard ratio; MAFLD, dysfunction-associated fatty liver disease; MASLD, metabolic dysfunction-associated steatotic liver disease; NAFLD, non-alcoholic fatty liver disease metabolic.

**Table S7. Association of advanced fibrosis status and all-cause mortality among individuals with NAFLD, MAFLD, MASLD and pure-MASLD (for the sensitivity analysis 1#).**

| | N | n (deaths) | Univariable model | | Multivariable Model 1 | | Multivariable Model 2 | |
|---|---|---|---|---|---|---|---|---|
| | | | HR (95% CI) | P | HR (95% CI) | P | HR (95% CI) | P |
| **NAFLD** | | | | | | | | |
| Low NFS | 1,310 | 298 | 1 | | 1 | | 1 | |
| Intermediate NFS | 548 | 333 | 4.17 (3.24, 5.35) | <0.001 | 1.24 (1.00, 1.55) | 0.052 | 1.27 (1.03, 1.57) | 0.025 |
| High NFS | 97 | 79 | 7.08 (4.49, 11.2) | <0.001 | 1.85 (1.23, 2.76) | 0.003 | 1.76 (1.21, 2.56) | 0.003 |
| **MAFLD** | | | | | | | | |
| Low NFS | 1,079 | 304 | 1 | | 1 | | 1 | |
| Intermediate NFS | 578 | 364 | 3.72 (2.91, 4.75) | <0.001 | 1.44 (1.16, 1.78) | <0.001 | 1.53 (1.24, 1.88) | <0.001 |
| High NFS | 104 | 85 | 5.96 (3.87, 9.18) | <0.001 | 1.89 (1.31, 2.74) | <0.001 | 1.79 (1.28, 2.51) | <0.001 |
| **MASLD** | | | | | | | | |
| Low NFS | 1,266 | 334 | 1 | | 1 | | 1 | |
| Intermediate NFS | 606 | 380 | 3.92 (3.08, 5.00) | <0.001 | 1.37 (1.11, 1.69) | 0.003 | 1.4 (1.15, 1.72) | <0.001 |
| High NFS | 107 | 86 | 6.03 (3.92, 9.27) | <0.001 | 1.76 (1.22, 2.54) | 0.002 | 1.61 (1.11, 2.32) | 0.012 |
| **Pure-MASLD** | | | | | | | | |
| Low NFS | 1,132 | 289 | 1 | | 1 | | 1 | |
| Intermediate NFS | 541 | 332 | 3.86 (3.05, 4.89) | <0.001 | 1.28 (1.02, 1.59) | 0.03 | 1.32 (1.06, 1.64) | 0.013 |
| High NFS | 95 | 78 | 6.81 (4.34, 10.7) | <0.001 | 1.84 (1.19, 2.84) | 0.006 | 1.76 (1.18, 2.62) | 0.006 |

### (n=6,966) including individuals with viral hepatitis or low BMI (<18.5 kg/m$^2$).

Survey-weight adjusted multivariable Cox proportional models were used in this analysis.

Multivariate model 1 was adjusted for sex, age, and race/ethnicity.

Multivariate model 2 was further adjusted for marital status, education, sedentary lifestyle, and smoking status in addition to model 1.

CI, confidential intervals; HR, hazard ratio; SLD, steatosis liver disease; MAFLD, dysfunction-associated fatty liver disease; MASLD, metabolic dysfunction-associated steatotic liver disease; NAFLD, non-alcoholic fatty liver disease metabolic; NFS, NAFLD fibrosis score.

**Table S8. Association SLD and all-cause mortality in individuals with different extent of hepatic steatosis.**

| | N | n (deaths) | Univariable model | | Multivariable Model 1 | | Multivariable Model 2 | |
|---|---|---|---|---|---|---|---|---|
| | | | HR (95% CI) | P | HR (95% CI) | P | HR (95% CI) | P |
| No hepatic steatosis | 4,544 | 1,244 | 1 | | 1 | | 1 | |
| Mild SLD | 871 | 251 | 1.05 (0.87, 1.27) | 0.6 | 0.97 (0.78, 1.21) | 0.8 | 0.95 (0.77, 1.18) | 0.7 |
| Moderate SLD | 902 | 383 | 1.99 (1.70, 2.32) | <0.001 | 1.23 (1.08, 1.39) | 0.001 | 1.21 (1.06, 1.39) | 0.005 |
| Severe SLD | 391 | 183 | 2.41 (1.77, 3.28) | <0.001 | 1.21 (1.01, 1.46) | 0.039 | 1.22 (1.03, 1.45) | 0.021 |

Survey-weight adjusted multivariable Cox proportional models were used in this analysis.

Multivariate model 1 was adjusted for sex, age, and race/ethnicity.

Multivariate model 2 was further adjusted for marital status, education, sedentary lifestyle, and smoking status in addition to model 1.

CI, confidential intervals; HR, hazard ratio; SLD, steatosis liver disease.

**Table S9. Association different subgroups of SLD and all-cause mortality according to the definition of MASLD (for the sensitivity analysis 2#).**

| | N | n (deaths) | Univariable model | | Multivariable Model 1 | | Multivariable Model 2 | |
|---|---|---|---|---|---|---|---|---|
| | | | HR (95% CI) | P | HR (95% CI) | P | HR (95% CI) | P |
| No hepatic steatosis | 4,544 | 1,244 | 1 | | 1 | | 1 | |
| Pure_MASLD | 1,092 | 486 | 2.15 (1.88, 2.46) | <0.001 | 1.2 (1.08, 1.34) | <0.001 | 1.21 (1.07, 1.35) | 0.001 |
| MetALD | 101 | 55 | 3.22 (2.31, 4.49) | <0.001 | 1.51 (1.00, 2.26) | 0.049 | 1.41 (1.02, 1.94) | 0.037 |
| MASLD_ALD | 30 | 18 | 2.28 (0.64, 8.05) | 0.2 | 1.95 (0.93, 4.08) | 0.075 | 1.95 (0.97, 3.91) | 0.061 |
| Steatosis without MASLD | 70 | 7 | 0.38 (0.13, 1.14) | 0.084 | 0.64 (0.23, 1.77) | 0.4 | 0.64 (0.26, 1.57) | 0.3 |

### (n=5,837) excluding individuals with mild hepatic steatosis from the SLD.

Survey-weight adjusted multivariable Cox proportional models were used in this analysis.

Multivariate model 1 was adjusted for sex, age, and race/ethnicity.

Multivariate model 2 was further adjusted for marital status, education, sedentary lifestyle, and smoking status in addition to model 1.

ALD, alcohol-related liver disease; CI, confidential intervals; HR, hazard ratio; SLD, steatosis liver disease; MASLD, metabolic dysfunction-associated steatotic liver disease.

**Table S10. Association among NAFLD, MAFLD, MASLD and pure-MASLD status, and cardiovascular disease and cancer-related mortality (for the sensitivity analysis 2[#]).**

| | | | | N | n (deaths) | Univariable model HR (95% CI) | Univariable model P | Multivariable Model 1 HR (95% CI) | Multivariable Model 1 P | Multivariable Model 2 HR (95% CI) | Multivariable Model 2 P |
|---|---|---|---|---|---|---|---|---|---|---|---|
| **Cardiovascular mortality** | | | | | | | | | | | |
| No NAFLD | | | | 4,682 | 340 | 1 | | 1 | | 1 | |
| NAFLD | | | | 1,155 | 136 | 1.96 (1.36, 2.83) | <0.001 | 1.07 (0.77, 1.49) | 0.7 | 1.08 (0.77, 1.52) | 0.7 |
| No MAFLD | | | | 4,700 | 329 | 1 | | 1 | | 1 | |
| MAFLD | | | | 1,137 | 147 | 2.44 (1.85, 3.22) | <0.001 | 1.18 (0.90, 1.54) | 0.2 | 1.19 (0.91, 1.57) | 0.2 |
| No MASLD | | | | 4,614 | 322 | 1 | | 1 | | 1 | |
| MASLD | | | | 1,223 | 154 | 2.24 (1.64, 3.05) | <0.001 | 1.14 (0.86, 1.51) | 0.4 | 1.14 (0.85, 1.53) | 0.4 |
| No pure-MASLD | | | | 4,745 | 341 | 1 | | 1 | | 1 | |
| Pure-MASLD | | | | 1,092 | 135 | 2.11 (1.47, 3.02) | <0.001 | 1.1 (0.79, 1.53) | 0.6 | 1.11 (0.78, 1.56) | 0.6 |
| No hepatic steatosis | | | | 4,544 | 319 | 1 | | 1 | | 1 | |
| NAFLD | MAFLD | MASLD | pure-MASLD | | | | | | | | |
| + | + | + | + | 1,022 | 130 | 2.37 (1.70, 3.29) | <0.001 | 1.15 (0.83, 1.58) | 0.4 | 1.16 (0.84, 1.62) | 0.4 |
| - | + | + | - | 115 | 17 | 2.75 (1.39, 5.41) | 0.003 | 1.33 (0.63, 2.82) | 0.5 | 1.27 (0.66, 2.46) | 0.5 |
| + | - | + | + | 70 | 5 | 0.48 (0.09, 2.67) | 0.4 | 0.53 (0.11, 2.47) | 0.4 | 0.42 (0.07, 2.37) | 0.3 |
| + | - | - | - | 63 | 1 | 0.01 (0.00, 0.10) | <0.001 | 0.02 (0.00, 0.17) | <0.001 | 0.02 (0.00, 0.18) | <0.001 |
| - | - | + | - | 16 | 2 | 2.13 (0.41, 11.0) | 0.4 | 1.04 (0.12, 8.79) | >0.9 | 1.22 (0.21, 7.02) | 0.8 |
| - | - | - | - | 7 | 2 | 2.71 (0.33, 22.0) | 0.4 | 12.2 (2.22, 67.4) | 0.004 | 6.82 (1.14, 40.6) | 0.035 |
| **Cancer mortality** | | | | | | | | | | | |
| No NAFLD | | | | 4,682 | 349 | 1 | | 1 | | 1 | |
| NAFLD | | | | 1,155 | 114 | 1.59 (1.18, 2.15) | 0.003 | 0.99 (0.74, 1.33) | >0.9 | 1.08 (0.81, 1.44) | 0.6 |
| No MAFLD | | | | 4,700 | 342 | 1 | | 1 | | 1 | |
| MAFLD | | | | 1,137 | 121 | 2.02 (1.55, 2.64) | <0.001 | 1.12 (0.85, 1.48) | 0.4 | 1.2 (0.91, 1.58) | 0.2 |

| | | | | | | | | | | | |
|---|---|---|---|---|---|---|---|---|---|---|---|
| No MASLD | | | | 4,614 | 333 | 1 | | 1 | | 1 | |
| MASLD | | | | 1,223 | 130 | 1.88 (1.44, 2.46) | <0.001 | 1.1 (0.84, 1.44) | 0.5 | 1.17 (0.89, 1.54) | 0.3 |
| No pure-MASLD | | | | 4,745 | 351 | 1 | | 1 | | 1 | |
| Pure-MASLD | | | | 1,092 | 112 | 1.64 (1.19, 2.25) | 0.002 | 0.99 (0.72, 1.35) | >0.9 | 1.07 (0.78, 1.46) | 0.7 |
| No hepatic steatosis | | | | 4,544 | 331 | 1 | | 1 | | 1 | |
| NAFLD | MAFLD | MASLD | pure-MASLD | | | | | | | | |
| + | + | + | + | 1,022 | 106 | 1.86 (1.36, 2.53) | <0.001 | 1.05 (0.77, 1.43) | 0.8 | 1.14 (0.83, 1.55) | 0.4 |
| - | + | + | - | 115 | 15 | 3.16 (1.59, 6.29) | 0.001 | 1.65 (0.92, 2.97) | 0.1 | 1.54 (0.84, 2.83) | 0.2 |
| + | - | + | + | 70 | 6 | 0.43 (0.13, 1.46) | 0.2 | 0.5 (0.15, 1.73) | 0.3 | 0.51 (0.14, 1.85) | 0.3 |
| + | - | - | - | 63 | 2 | 0.89 (0.21, 3.72) | 0.9 | 1.21 (0.33, 4.42) | 0.8 | 1.35 (0.44, 4.14) | 0.6 |
| - | - | + | - | 16 | 3 | 3.67 (0.91, 14.8) | 0.068 | 2.38 (0.38, 15.1) | 0.4 | 2.34 (0.42, 13.0) | 0.3 |
| - | - | - | - | 7 | 0 | / | | / | | / | |

[#] (n=5,837) excluding individuals with mild hepatic steatosis from the SLD.

Survey-weight adjusted multivariable Cox proportional models were used in this analysis.

Multivariate model 1 was adjusted for sex, age, and race/ethnicity.

Multivariate model 2 was further adjusted for marital status, education, sedentary lifestyle, and smoking status in addition to model 1.

CI, confidential intervals; HR, hazard ratio; SLD, steatosis liver disease; MAFLD, dysfunction-associated fatty liver disease; MASLD, metabolic dysfunction-associated steatotic liver disease; NAFLD, non-alcoholic fatty liver disease metabolic.

**Table S11. Association of advanced fibrosis status and all-cause mortality among individuals with NAFLD, MAFLD, MASLD and pure-MASLD (for the sensitivity analysis 2#).**

| | N | n (deaths) | Univariable model | | Multivariable Model 1 | | Multivariable Model 2 | |
|---|---|---|---|---|---|---|---|---|
| | | | HR (95% CI) | P | HR (95% CI) | P | HR (95% CI) | P |
| **NAFLD** | | | | | | | | |
| Low NFS | 701 | 193 | 1 | | 1 | | 1 | |
| Intermediate NFS | 378 | 240 | 3.84 (2.81, 5.24) | <0.001 | 1.41 (1.09, 1.83) | 0.009 | 1.55 (1.19, 2.01) | 0.001 |
| High NFS | 62 | 50 | 6.27 (3.93, 10.0) | <0.001 | 1.79 (1.02, 3.11) | 0.041 | 1.82 (1.09, 3.03) | 0.022 |
| **MAFLD** | | | | | | | | |
| Low NFS | 650 | 204 | 1 | | 1 | | 1 | |
| Intermediate NFS | 405 | 262 | 3.47 (2.55, 4.73) | <0.001 | 1.51 (1.16, 1.95) | 0.002 | 1.6 (1.22, 2.09) | <0.001 |
| High NFS | 68 | 55 | 5.57 (3.36, 9.23) | <0.001 | 1.82 (1.12, 2.93) | 0.015 | 1.89 (1.20, 3.00) | 0.006 |
| **MASLD** | | | | | | | | |
| Low NFS | 719 | 220 | 1 | | 1 | | 1 | |
| Intermediate NFS | 419 | 274 | 3.66 (2.71, 4.96) | <0.001 | 1.49 (1.17, 1.89) | 0.001 | 1.6 (1.26, 2.04) | <0.001 |
| High NFS | 69 | 56 | 5.88 (3.62, 9.54) | <0.001 | 1.81 (1.13, 2.88) | 0.013 | 1.87 (1.19, 2.94) | 0.006 |
| **Pure-MASLD** | | | | | | | | |
| Low NFS | 645 | 190 | 1 | | 1 | | 1 | |
| Intermediate NFS | 373 | 249 | 3.71 (2.71, 5.07) | <0.001 | 1.44 (1.11, 1.87) | 0.006 | 1.57 (1.21, 2.04) | <0.001 |
| High NFS | 61 | 50 | 6.73 (4.19, 10.8) | <0.001 | 1.84 (1.05, 3.22) | 0.034 | 1.85 (1.10, 3.12) | 0.021 |

### (n=5,837) excluding individuals with mild hepatic steatosis from the SLD.

Survey-weight adjusted multivariable Cox proportional models were used in this analysis.

Multivariate model 1 was adjusted for sex, age, and race/ethnicity.

Multivariate model 2 was further adjusted for marital status, education, sedentary lifestyle, and smoking status in addition to model 1.

CI, confidential intervals; HR, hazard ratio; SLD, steatosis liver disease; MAFLD, dysfunction-associated fatty liver disease; MASLD, metabolic dysfunction-associated steatotic liver disease; NAFLD, non-alcoholic fatty liver disease metabolic; NFS, NAFLD fibrosis score.
